# Publication bias in pharmacogenetics of statin-associated muscle symptoms, an umbrella review with a meta-epidemiological study

**DOI:** 10.1101/2024.03.06.24303892

**Authors:** A Gougeon, I Aribi, S Guernouche, JC Lega, JM Wright, C Verstuyft, A Lajoinie, F Gueyffier, G Grenet

## Abstract

**Background:** Statin-associated muscle symptoms (SAMS) are a major cause of treatment discontinuation. Adjusting statin dosages for solute carrier organic anion transporter family member 1B1 (SLCO1B1) genotype has been proposed to reduce SAMS. We hypothesized that the association between SLCO1B1 genotype and SAMS is misestimated because of publication bias.

**Methods:** We searched for published systematic reviews evaluating the association between SLCO1B1 genotype and SAMS. We collected the odds ratio (OR) of this association in each clinical study. We assessed the presence of publication bias using the visual inspection of a funnel plot and Egger’s test and used the Bayes Factor (BF_Publication-bias_) of the Robust Bayesian Meta-Analysis (RoBMA) as a sensitivity analysis. We evaluated the effect of publication bias by comparing qualitatively and quantitatively (ratio of OR [ROR]) OR of the meta-analysis i) uncorrected for potential publication bias (OR_Uncorrected_) and ii) corrected using the trim-and-fill (OR_Trim&Fill_). We also used the RoBMA (OR_RoBMA_) for corrected OR as a sensitivity analysis. Our primary analysis covered the associations between any SLCO1B1 genotype and any statin drug. Secondary analysis focused on SLCO1B1 genotypes and statin drug subgroups.

**Results:** We included 8 cohort and 11 case-control studies, totaling 62 OR of three SLCO1B1 genotypes and five statin drugs plus one ‘mixed’ statin treatment. All controls were statin-tolerant patients. In the primary analysis, the funnel plot was suggestive of publication bias, confirmed by Egger’s test (p=0.001) and RoBMA (BF_Publication-bias_=18). Correcting the estimate for publication bias resulted in loss of the association, from a significant OR_Uncorrected_ (1.31 95% CI [1.13– 1.53]) to corrected ORs suggesting no difference: i) OR_Trim&Fill_ (1.07 95% CI [0.89–1.30]) and ii) OR_RoBMA_ (1.02 95% CI [1.00–1.33]). The ROR_Trim&Fill_ and the ROR_RoBMA_ suggested that publication bias overestimated the association by 18% and 23%, respectively. The results were similar for the most studied SLCO1B1 genotype, as for simvastatin and atorvastatin.

**Conclusion:** The effect of the SLCO1B1 genotype on the risk of developing SAMS is overestimated in the published literature. This could lead prescribers to incorrectly decreasing statin doses or even avoiding statin use, leading to a loss of the potential cardiovascular benefit of statins.

**Clinical perspective:** What is new?

- There is significant publication bias in the available literature regarding the association between SLCO1B1 genotype and statin-associated muscle symptoms.
- The available literature overestimates the importance of the SLCO1B1 genotype on statin-associated muscle symptoms.

What are the clinical implications?

- The cardiovascular benefit of statins might be wrongly lost when adjusting statin therapy with the SLCO1B1 genotype.
- The effect of publication bias should be considered when writing guidelines.

## INTRODUCTION

Statins decrease low-density lipoprotein cholesterol (LDL-C). Several randomized controlled trials (RCTs) have confirmed the cardiovascular (CV) benefits of several statins in different primary and secondary prevention populations [1–3]. The most common reported adverse events (AEs) are statin-associated muscle symptoms (SAMS). SAMS range from myalgia to rhabdomyolysis, with or without elevated creatine kinase (CK); they decrease or stop after treatment discontinuation [4]. The exact mechanism of statin muscle toxicity has yet to be fully clarified [5]. SAMS occurrence is likely to limit patient adherence to statins. The prevalence of muscle symptoms in treated patients has been estimated to be around 10% [range 5%–25%]. More specifically, muscular symptoms only attributable to statins have been estimated around 1-2% [ranging from 0.5% to 4%] [4]. Several genotypes of the solute carrier organic anion transporter family member 1B1 (SLCO1B1) gene have been reported to be associated with an increased risk of SAMS [6–11]. The SLCO1B1 gene encodes for the OATP1B1 protein, which is involved in the hepatic transport of statins. The European Medicines Agency (EMA) and the Food and Drug Administration (FDA) have listed the SLCO1B1 gene mutation as a potential marker for SAMS. The Clinical Pharmacogenetics Implementation Consortium (CPIC) recently updated their guidelines and recommend dose adjustment for statin treatment according to SLCO1B1 genotype “if pharmacogenetic test results are available” to reduce the risk of SAMS [12]. The impact of genetic information on prescription changes according to CPIC guidelines was evaluated in a recent study [13]. In 64% of cases, decision aids were used by prescribers, resulting in: a reduced dose of simvastatin in 74% and change in the statin in 26%. Although CPIC does not endorse genetic testing to determine the SLCO1B1 genotype, their estimates suggest that 11-36% of individuals have an intermediate activity phenotype and 0-6% have a low activity phenotype [12].

However, the supposed association between SAMS and SLCO1B1 genotype might be affected by publication bias [14]. Such publication bias has been documented in drug safety assessment [15], genetic epidemiology [16] and more recently in pharmacogenetics [17]. Regarding the SLCO1B1—SAMS association, the available systematic reviews suggest a potential significant publication bias but failed to take it into account. The meta-analysis of Jiang *et al.* reported a significant association between a SLCO1B1 genotype and an increased risk of AE for various statins [18]. However, they also reported significant Egger’s and Begg’s tests for one association, but without including this particular risk of bias in their conclusion or in the abstract. Moreover, they acknowledged in the discussion section that their review might lack power to detect publication bias. More recently, Xiang *et al.* reported a significant increased risk of SAMS with specific SLCO1B1 genotype [19]. However, among the eight Egger’s and Begg’s tests they reported, four were significant (p value <5%). Moreover, they used the trim-and-fill (T&F) correction to take into account the publication bias, and reported a loss of the association for one out the three reported corrected ORs [20]. However, they did not state this publication bias and its impact in the conclusion or in the abstract. The presence of this uncertainty poses a significant concern, considering the fact that guidelines are recommending the use of SLCO1B1 genotype to reduce the incidence of SAMS [12]. Consequently, accurately estimating the association between the SLCO1B1 genotype and SAMS is imperative for making well-informed clinical decisions.

Therefore, we aimed to systematically assess the presence and the effect of publication bias on the association between SLCO1B1 genotype and SAMS.

## METHODS

### Protocol and registration

The protocol was registered *a priori* on the Open Science Framework (OSF) website: https://osf.io/36jq8/. Our methodology complied with the Cochrane Handbook for Systematic Reviews of Interventions [21]. We reported our study in accordance with the Preferred Reporting Items for Systematic Reviews and Meta-Analyses (PRISMA) checklist (Supplemental material [S1]) [22]. The current study follows the methodology already used in a similar work we previously published [17].

### Search strategy and inclusion criteria

We conducted an umbrella review of the literature up to November 2022. We used PubMed, Embase and Cochrane Central. Our search terms were: “[polymorphism genetic OR solute carrier organic anion transporter 1B1 OR SLCO1B1 OR organic anion transporter polypeptide 1B1 OR OATP1B1] AND [statin OR rosuvastatin OR fluvastatin OR pravastatin OR simvastatin OR cerivastatin OR lovastatin OR atorvastatin OR pitavastatin] AND [myopathy OR myalgia OR myositis OR rhabdomyolysis OR creatine kinase OR CK]” (see S2 for details). We restricted the search to articles published in English. In addition, we performed a manual search in the references of articles obtained by the automatic search. Three reviewers (AG, IA, SG) independently selected the articles based on title and abstract then on full text. Any disagreement over article selection was resolved by consensus. Our PICOt criteria were as follow: (i) patient: humans, (ii) intervention: any statin, (iii) comparator: any control, (iv) outcome: point estimate of SLCO1B1-SAMS association, (v) type of publication: systematic reviews and meta-analyses. Our inclusion criteria were: (i) systematic reviews published in peer-reviewed journals [23], (ii) examining the association between SLCO1B1 genotype and SAMS, (iii) including clinical studies, (iv) published in English, and (v) calculation of point estimate using meta-analyses. We focused on English-published studies as they are more likely to be used in guidelines. Indeed, the CPIC guideline on the topic was restricted to English articles for the SLCO1B1 assessment [12]. In this way, our results are representative of the literature that is used for such guideline elaboration.

### Data extraction and quality assessment of included studies

Two independent reviewers (AG, IA) extracted the data using a standardized form. Extracted data included study characteristics (e.g. authors, year of publication), information on the statins studied (e.g. type, dose), genetic variants of the SLCO1B1 gene, muscle pain criteria, measures of association‒‒odds ratios (OR) and confidence intervals (CI), and adjustments for potential covariates. When some data of a clinical study were missing in the included systematic reviews, we extracted them from the original clinical study report. The risk of bias in the clinical studies was the one provided by the included systematic review, following the Cochrane Handbook recommendations for umbrella review [21].

### Dataset

Our dataset included all point estimates as OR and its 95% CI of the association between SAMS and SLCO1B1 genotype collected across the included systematic reviews. Clinical studies included in more than one systematic review were used only once (S3). The dataset also included characteristics associated with the point estimates: type of statin, type of polymorphism and genetic model (homozygous comparison, heterozygous comparison, dominant, allelic) used for subgroup analyses.

### Primary, secondary and exploratory analyses

Our primary analysis integrated all point estimates: any SLCO1B1 single nucleotide Polymorphism (SNPs), any statin. In the primary analysis, we prioritized the homozygous (CC vs TT) and heterozygous (TC vs TT) comparisons to avoid overlapping measurements for the same clinical study.

Our secondary analysis assessed publication bias in different subgroups: each SLCO1B1 SNP, focus each statin type and each genetic model (heterozygous comparison, homozygous comparison, allelic model - C vs T - or dominant model - TC + CC vs TT - ) when possible (≥ 10 point estimates needed for Egger’s test) [24]. We also performed an analysis focused on SNP rs4149056 because it is the most incriminating SNP in SAMS.

We also conducted exploratory analyses to look for a source of publication bias according to three factors: clinical study sponsors, statin drug names, and ethnic groups. For these three potential sources, we plotted the main funnel plot but distinguishing private versus public sponsors for the first one, statin drug names for the second one, and according to the ethnic groups as reported in the included systematic reviews for the third one, although the reporting of population differences in clinical research is inconsistent and plagued with methodological issues [25].

### Statistical analysis

We first used the funnel plot to assess the presence of publication bias. Two independent researchers (AG, IA) visually analyzed the funnel plots and classified them as: “suggestive of a publication bias”, “not suggestive of a publication bias”, or “not assessable”. Agreement was estimated using Free-marginal kappa estimator [26]. A third researcher (GG), blinded to the previous classifications, helped resolve disagreements. Second, we used Egger’s test for associations with at least 10 point estimates [24] . We did not use Begg’s test [27]. When the Egger’s test was significant, we added a sensitivity analysis using the Robust Bayesian Meta-Analysis (RoBMA) approach (see “sensitivity analysis” section) [28].

We estimated the SLCO1B1-SAMS association at the meta-analysis level i) without and ii) with a correction for publication bias. We used an inverse variance weighting random-effect meta-analysis to estimate the uncorrected OR (OR_Uncorrected_) and its 95% confidence interval (95% CI). Inter-study heterogeneity was assessed using the Cochran’s Q test, with significance set at p < 0.05, and the I^2^ was calculated [29]. When publication bias was confirmed by a significant p-value of the Egger’s test, we estimated a corrected OR (OR_Trim&Fill_) and its 95% CI, using the T&F method [20]. We then compared the estimate of the SLCO1B1-SAMS association i) qualitatively (significant or not significant association) and ii) quantitatively by calculating the ratio of the OR (OR_Trim&Fill_ / OR_Uncorrected_ ) and its 95% CI (ROR_Trim&Fill_) (see for details S4).

### Sensitivity analysis

We conducted sensitivity analyses using the recently proposed Robust Bayesian Meta-Analysis (RoBMA) approach [28]. RoBMA integrates several publication bias detection and correction methods, notably selection models that are less sensitive to heterogeneity [30]. RoBMA results use Bayes factors (BF), a continuous measure of evidence in favor of the presence or absence of effect, heterogeneity and publication bias. Bayes factor values above 10 indicate very strong evidence, from 3 to 10 moderate evidence, and from 1 to 3 weak evidence. In several simulation studies, RoBMA was found to be superior to other bias correction methods [28]. RoBMA has been used notably in the field of psychology [31–33]. We used RoBMA to provide a sensitivity analysis of i) the publication bias detection, when the Egger’s test was significant and ii) the evaluation of the publication bias effect, by providing a corrected OR_RoBMA_ and comparing it to the OR_Uncorrected_ as previously described with the T&F method. The ratio OR_RoBMA_ divided by OR_Uncorrected_ being ROR_RoBMA_.

We conducted the analyses using the R 4.2.2 software [34] and the package meta (6.2-1) [35] and RoBMA (2.3.1). All p-values were considered significant at 0.05 without multiple testing adjustment.

## RESULTS

### Study selection and characteristics

Out of 436 identified references, we included 6 systematic reviews, totaling 19 original clinical studies, from which we extracted 62 unique point estimates of the association between the SLCO1B1 genotype and SAMS (Figure 1).

**Figure 1:**
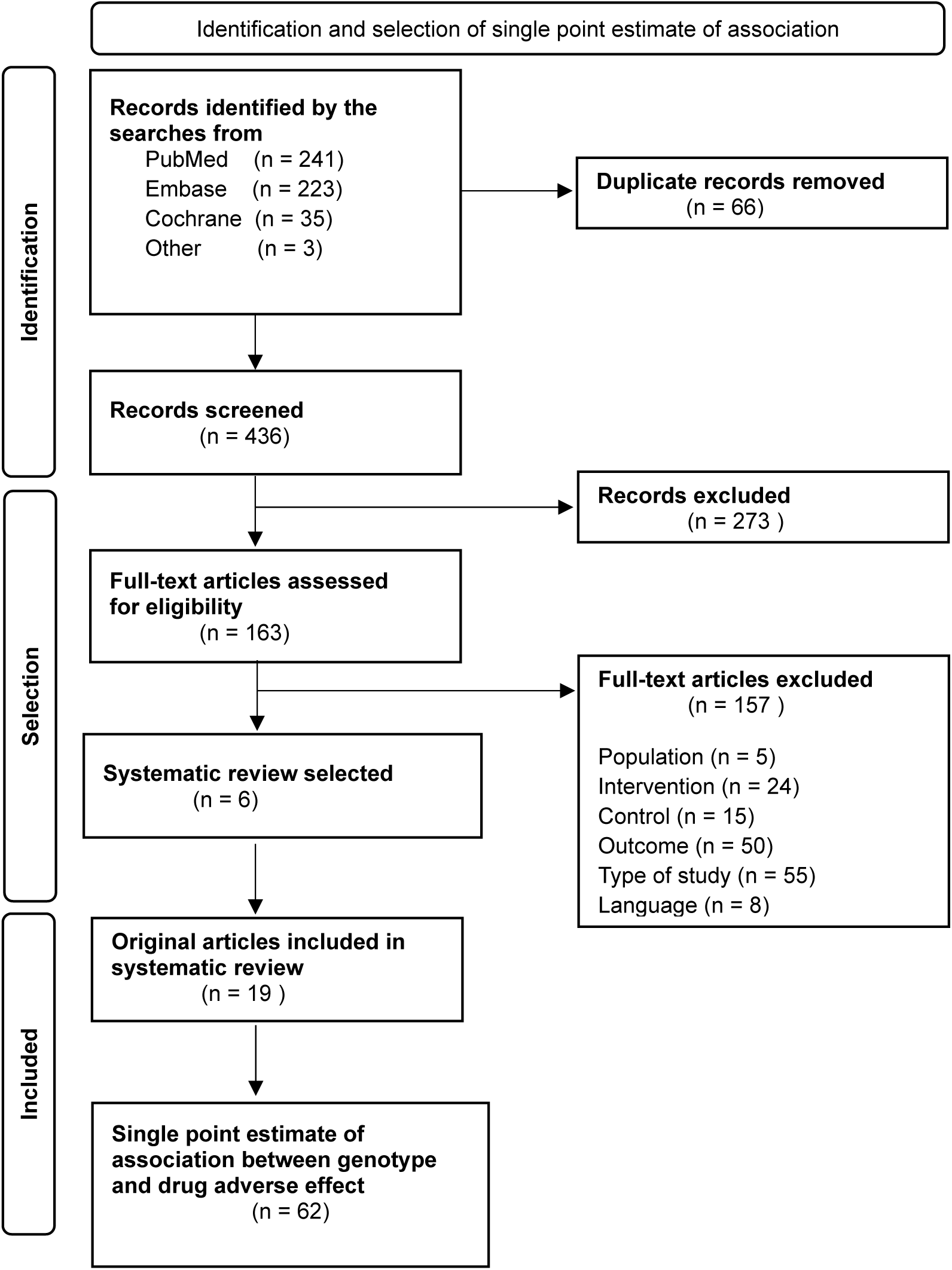
Flow chart diagram of bibliographic search.

All clinical studies used wild-type (WT) statin-tolerant patients as a control group, leading to an observational design for the assessment of this association for all studies (losing the benefit of randomization when they were part of a randomized trial). They evaluated one or more statins, mainly simvastatin and atorvastatin; 6 did not specify the statin drugs (statin mixed). Eighteen clinical studies evaluated rs4149056 single nucleotide polymorphisms (SNPs), 7 evaluated rs2306283 and 3 evaluated rs4363657 of the SLCO1B1 gene (Table 1). The criteria for assessing myopathy varied between studies, with the main criterion being muscle symptoms, irrespective of elevated CK levels. However, the reporting of the muscle adverse event was highly heterogeneous, precluding any subgroup constitution. The study population was predominantly of European descent. One clinical study was sponsored by a pharmaceutical company.

**Table 1:**
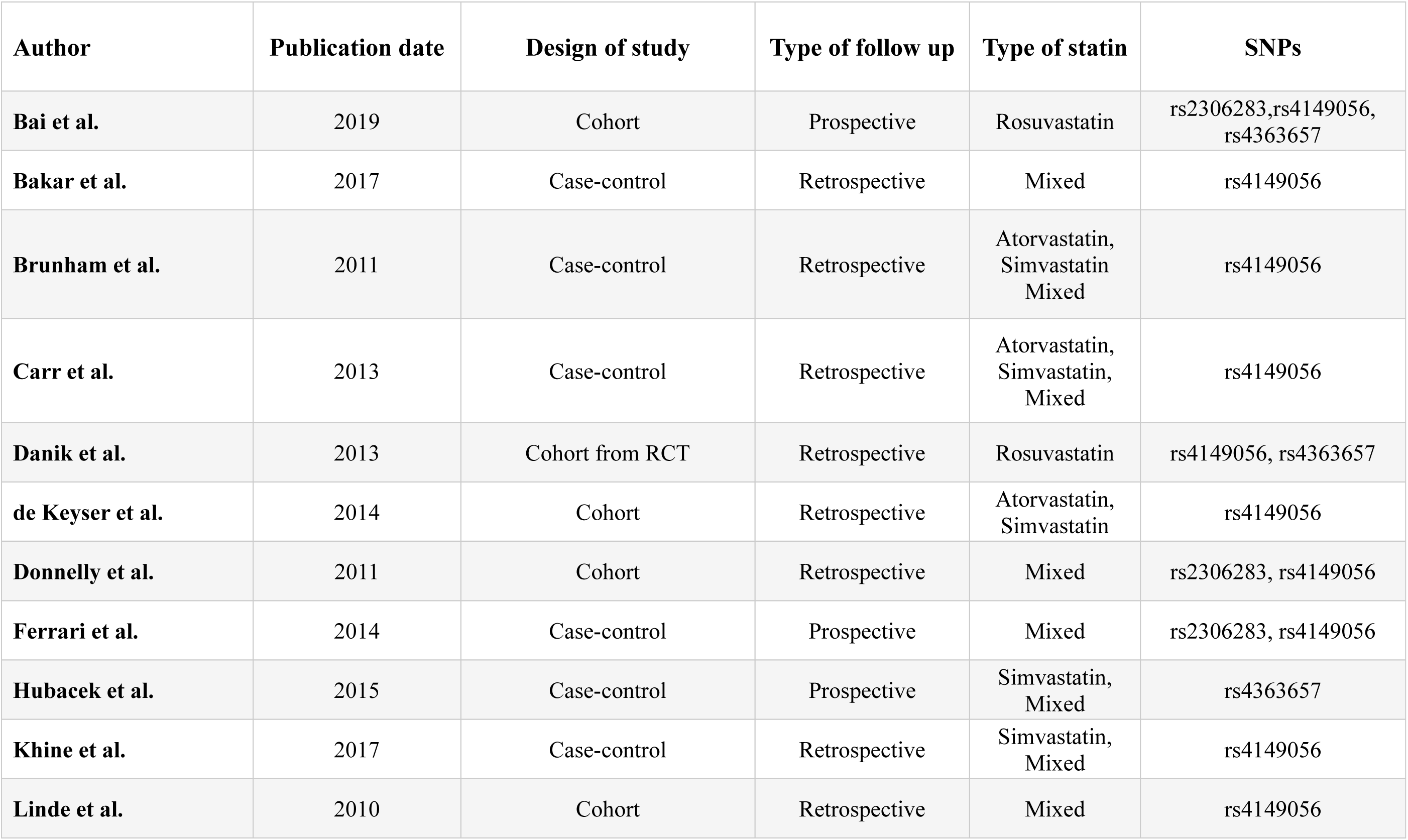
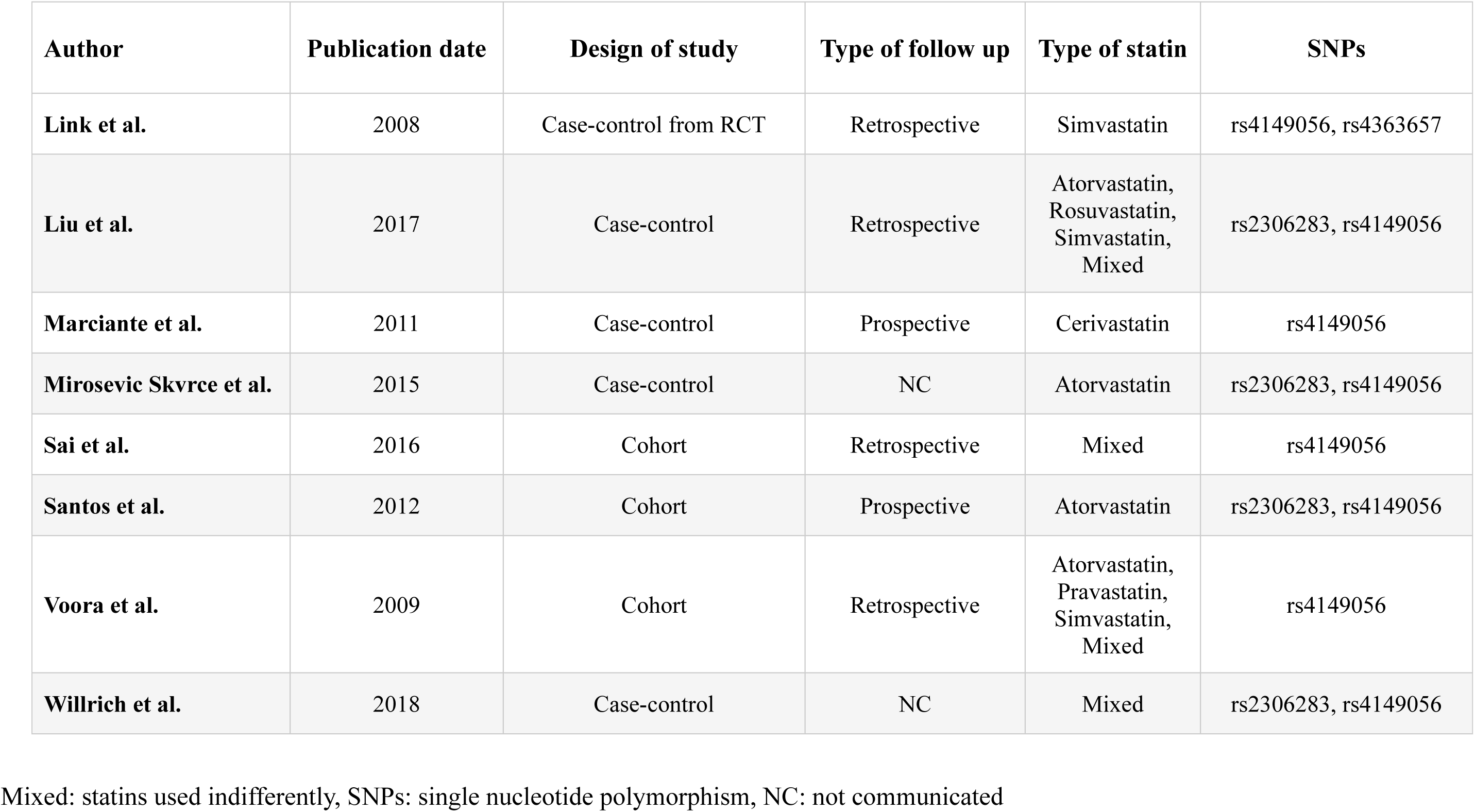
Characteristics of the clinical studies collected through the included systematic reviews.

| Author | Publication date | Design of study | Type of follow up | Type of statin | SNPs |
| --- | --- | --- | --- | --- | --- |
| Bai et al. | 2019 | Cohort | Prospective | Rosuvastatin | rs2306283,rs4149056,<br>rs4363657 |
| Bakar et al. | 2017 | Case-control | Retrospective | Mixed | rs4149056 |
| Brunham et al. | 2011 | Case-control | Retrospective | Atorvastatin,<br>Simvastatin<br>Mixed | rs4149056 |
| Carr et al. | 2013 | Case-control | Retrospective | Atorvastatin,<br>Simvastatin,<br>Mixed | rs4149056 |
| Danik et al. | 2013 | Cohort from RCT | Retrospective | Rosuvastatin | rs4149056, rs4363657 |
| de Keyser et al. | 2014 | Cohort | Retrospective | Atorvastatin,<br>Simvastatin | rs4149056 |
| Donnelly et al. | 2011 | Cohort | Retrospective | Mixed | rs2306283, rs4149056 |
| Ferrari et al. | 2014 | Case-control | Prospective | Mixed | rs2306283, rs4149056 |
| Hubacek et al. | 2015 | Case-control | Prospective | Simvastatin,<br>Mixed | rs4363657 |
| Khine et al. | 2017 | Case-control | Retrospective | Simvastatin,<br>Mixed | rs4149056 |
| Linde et al. | 2010 | Cohort | Retrospective | Mixed | rs4149056 |

| **Author** | **Publication date** | **Design of study** | **Type of follow up** | **Type of statin** | **SNPs** |
| --- | --- | --- | --- | --- | --- |
| **Link et al.** | 2008 | Case-control from RCT | Retrospective | Simvastatin | rs4149056, rs4363657 |
| **Liu et al.** | 2017 | Case-control | Retrospective | Atorvastatin,<br>Rosuvastatin,<br>Simvastatin,<br>Mixed | rs2306283, rs4149056 |
| **Marciante et al.** | 2011 | Case-control | Prospective | Cerivastatin | rs4149056 |
| **Mirosevic Skvrce et al.** | 2015 | Case-control | NC | Atorvastatin | rs2306283, rs4149056 |
| **Sai et al.** | 2016 | Cohort | Retrospective | Mixed | rs4149056 |
| **Santos et al.** | 2012 | Cohort | Prospective | Atorvastatin | rs2306283, rs4149056 |
| **Voora et al.** | 2009 | Cohort | Retrospective | Atorvastatin,<br>Pravastatin,<br>Simvastatin,<br>Mixed | rs4149056 |
| **Willrich et al.** | 2018 | Case-control | NC | Mixed | rs2306283, rs4149056 |

Mixed: statins used indifferently, SNPs: single nucleotide polymorphism, NC: not communicated

The risk of bias of the clinical studies was assessed using the Newcastle-Ottawa Scale (NOS) in 5 reviews and the Risk of Bias Score for Genetic Association Studies in 1 review (i.e. Turongkaravee, 2021). The overall risk of bias was low as all the point estimates of the SLCO1B1-SAMS associations were assessed using non-randomized comparisons (for details see S3,S5).

### Primary analysis

#### Detection of the presence of publication bias

We included 62 point estimates in the primary analysis. The visual inspection of the funnel plots was suggestive of publication bias (kappa agreement between the two reviewers: 64%). The visual inspection also revealed a possible clustering of the estimates around the significant p-value thresholds (Figure 2). The asymmetry was confirmed by Egger’s test (p=0.001) (Table 2). The sensitivity analysis with RoBMA showed strong evidence for publication bias with a BF_Publication-bias_ of 18 (Table 2, S6).

**Figure 2:**
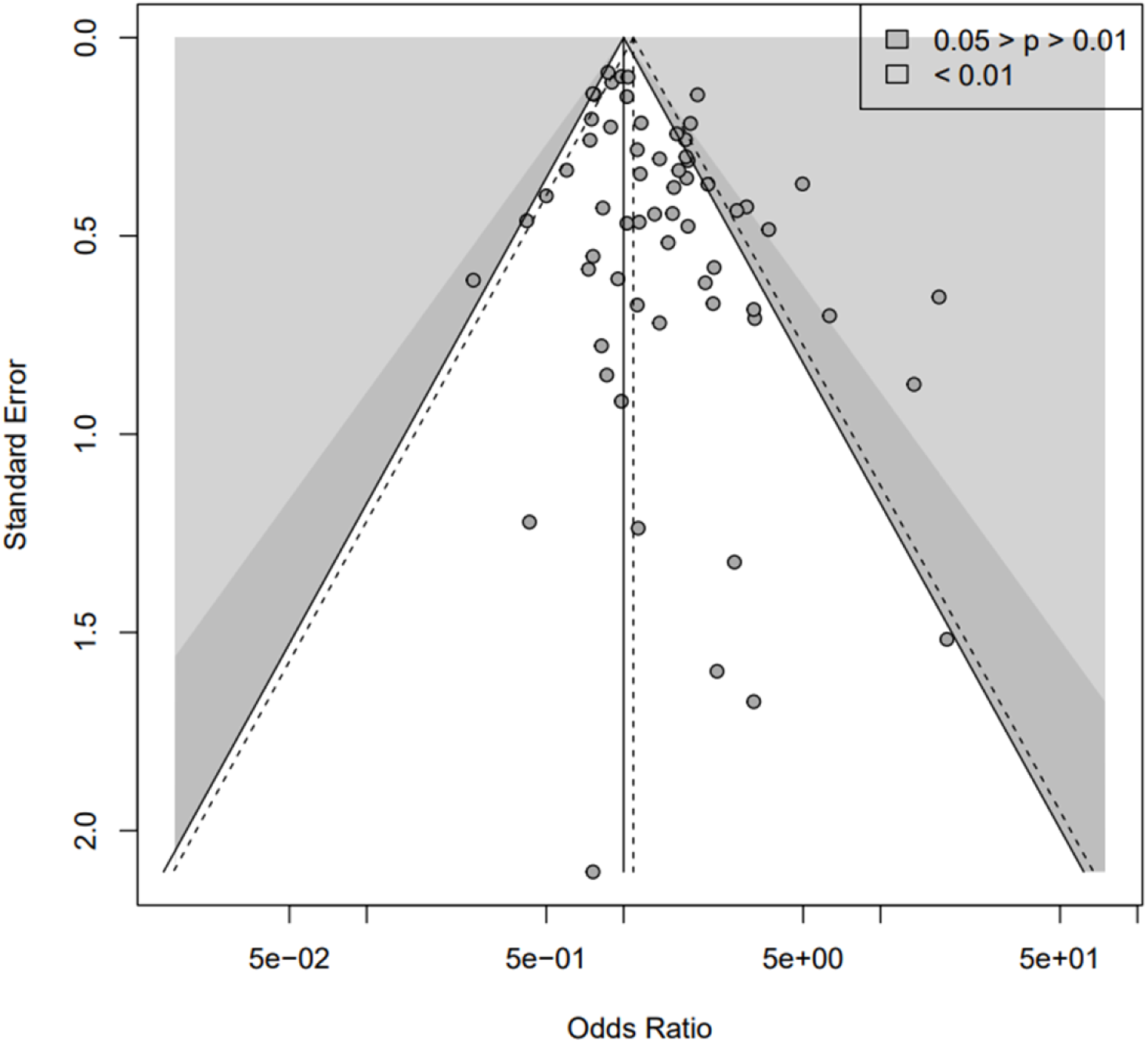
Funnel plot of the primary analysis (k = 62) [SLCO1B1 polymorphisms – Statins – SAMS]. - Funnel plot of the association between [SLCO1B1 – Statin – SAMS] k: number of point estimates. In the funnel plot, each point is an estimation of the associations. The white, dark and light grey zones stand for a p-value of the odds ratio i) non-significant, ii) between 0.05 and 0.01, and iii) <0.01, respectively. The dashed triangle stands for the estimation of the meta-analysis of the association, without adjusting for a potential publication bias.

**Table 2:**
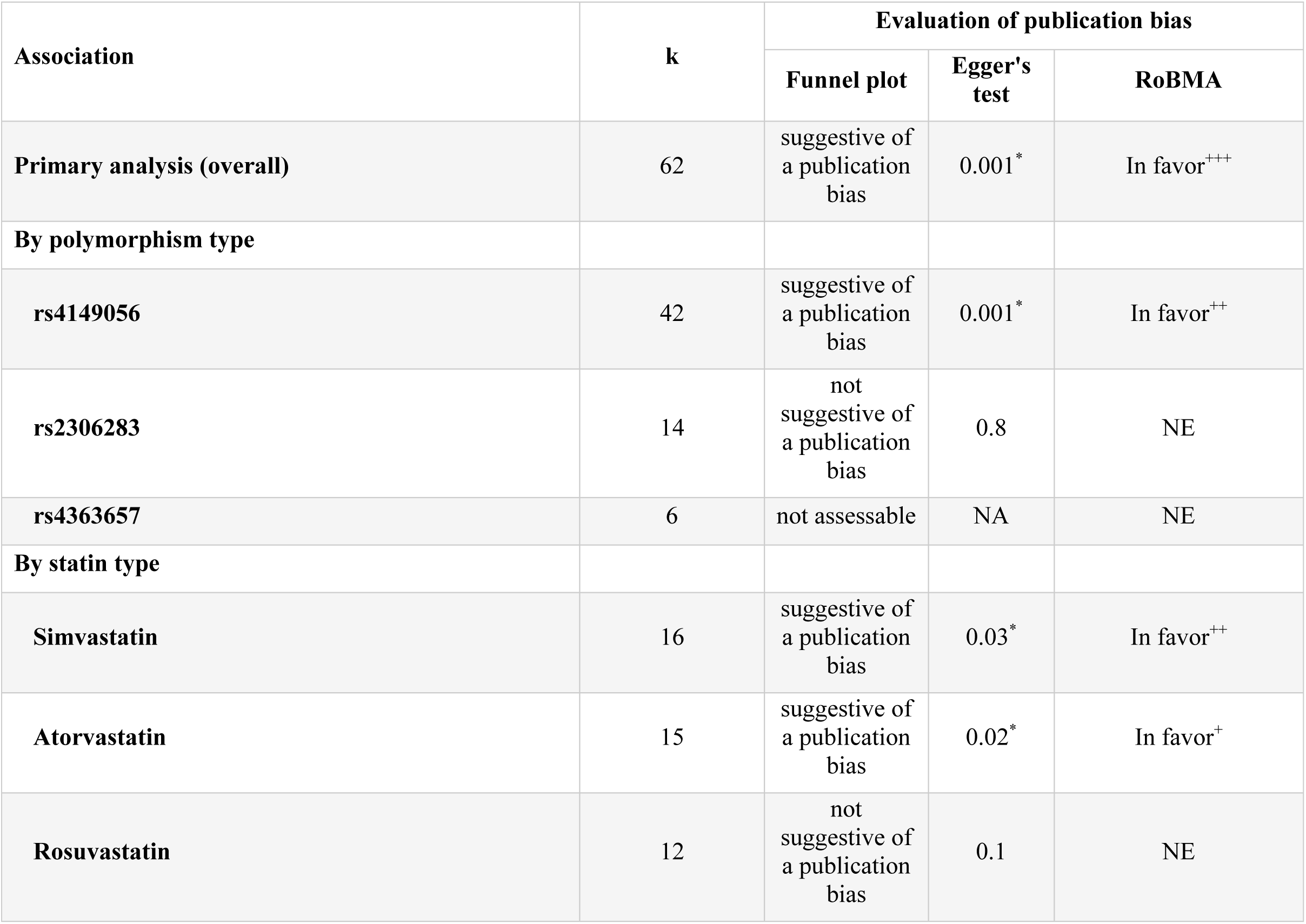
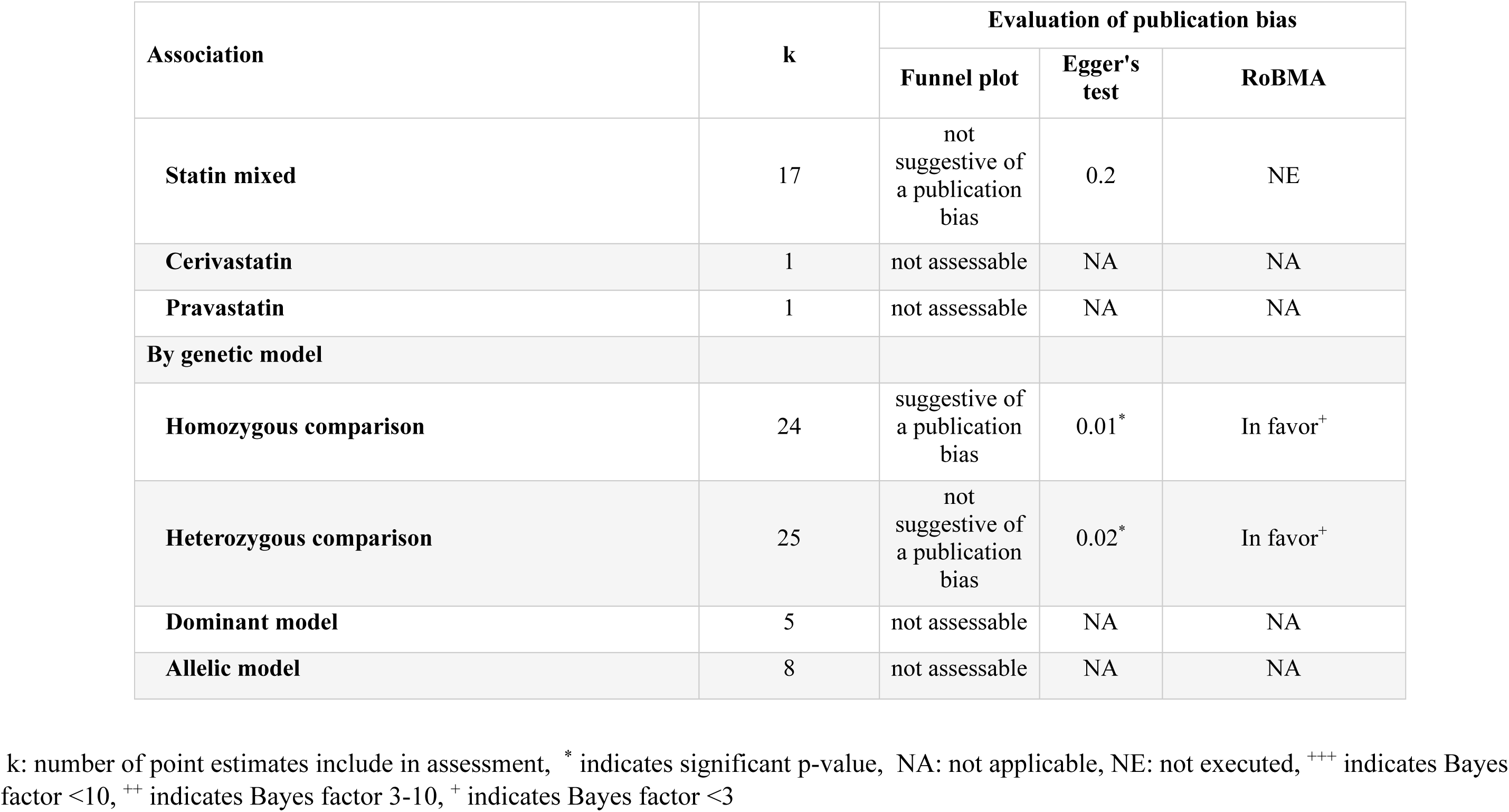
Detection of publication bias in the primary and secondary analysis.

#### Evaluation of the impact of publication bias on the association estimate

The uncorrected meta-analysis yielded an estimate with a 31% significant increased risk of SAMS with SLCO1B1 genotype (OR_Uncorrected_ = 1.31 CI 95% 1.13–1.53), with substantial heterogeneity (Cochran’s Q test p<0.05, I^2^=64%) (S7). Conversely, the corrected meta-analysis suggested no effect (OR_Trim&Fill_ = 1.07, CI 95% [0.89–1.30]). Correcting for publication bias led to a loss of the association and a 18% nonsignificant change in the association estimate (ROR_Trim&Fill_= 0.82, CI 95% 0.64–1.04) (Table 3).

**Table 3:**
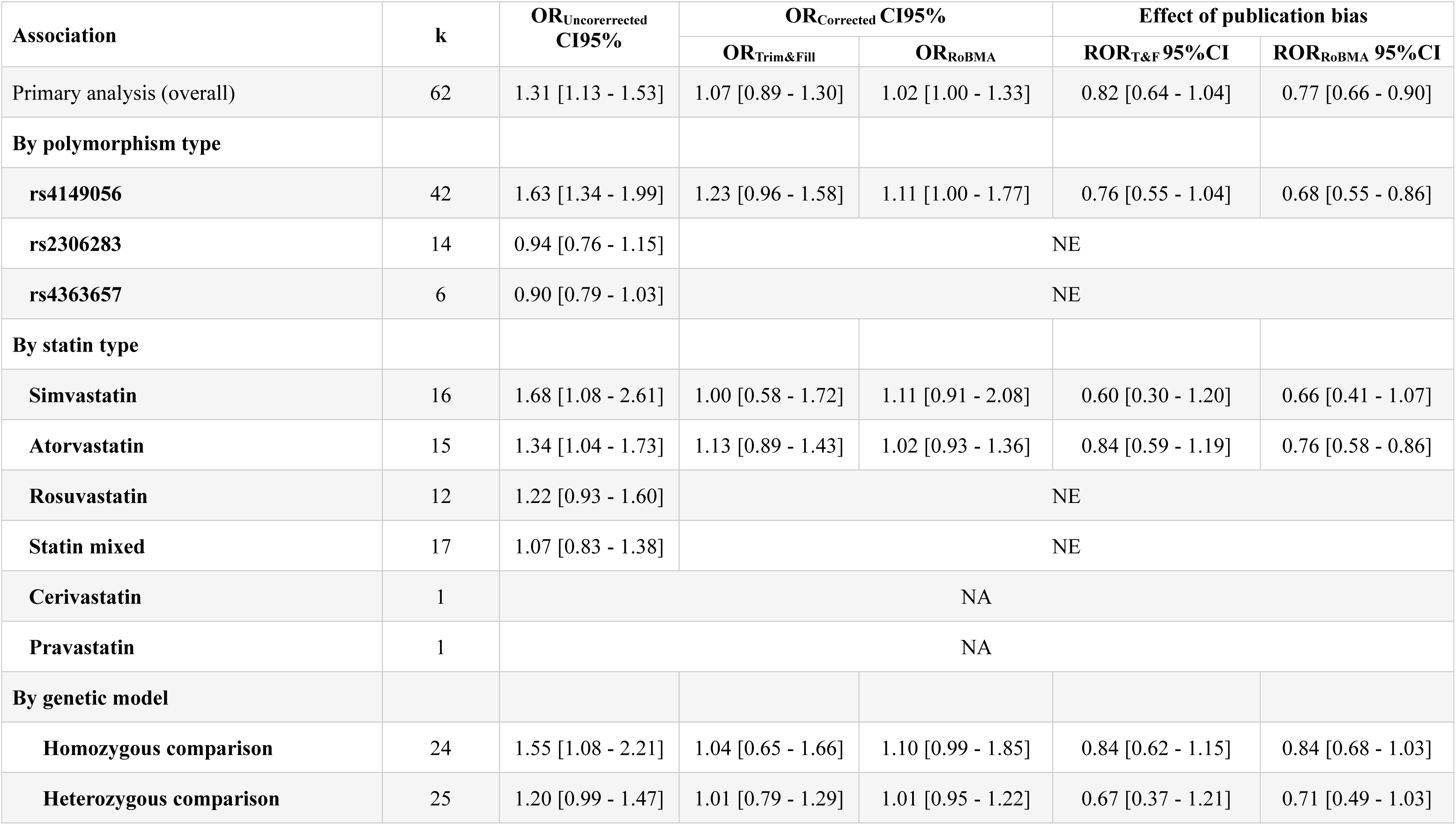
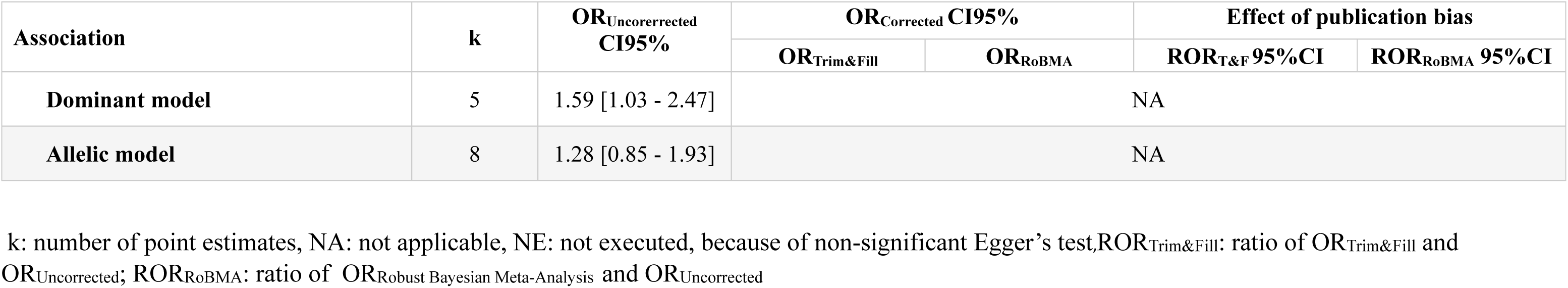
Estimate and correction of publication bias in the primary and secondary analysis.

The corrected meta-analysis using RoBMA also suggested no effect (OR_RoBMA_=1.02, CI 95% 1.00–1.33) in the sensitivity analysis. Correcting for publication bias resulted in a loss of the observed association, with a 23% significant change in the association estimate (ROR_RoBMA_=0.77, CI 95% 0.55–0.86) (Table 3).

### Secondary analysis

#### Detection of the presence of publication bias by statin type

We examined subgroups by statin type by incorporating 16 point estimates for simvastatin, 15 for atorvastatin, and 12 for rosuvastatin. The visual inspection of funnel plots and Egger’s tests suggested the presence of publication bias for simvastatin and atorvastatin (Figure 3 and Table 3). However, the visual inspection for atorvastatin revealed an asymmetry caused by non-significant point estimates. For rosuvastatin, the funnel plot and Egger’s test were not suggestive of publication bias.

**Figure 3:**
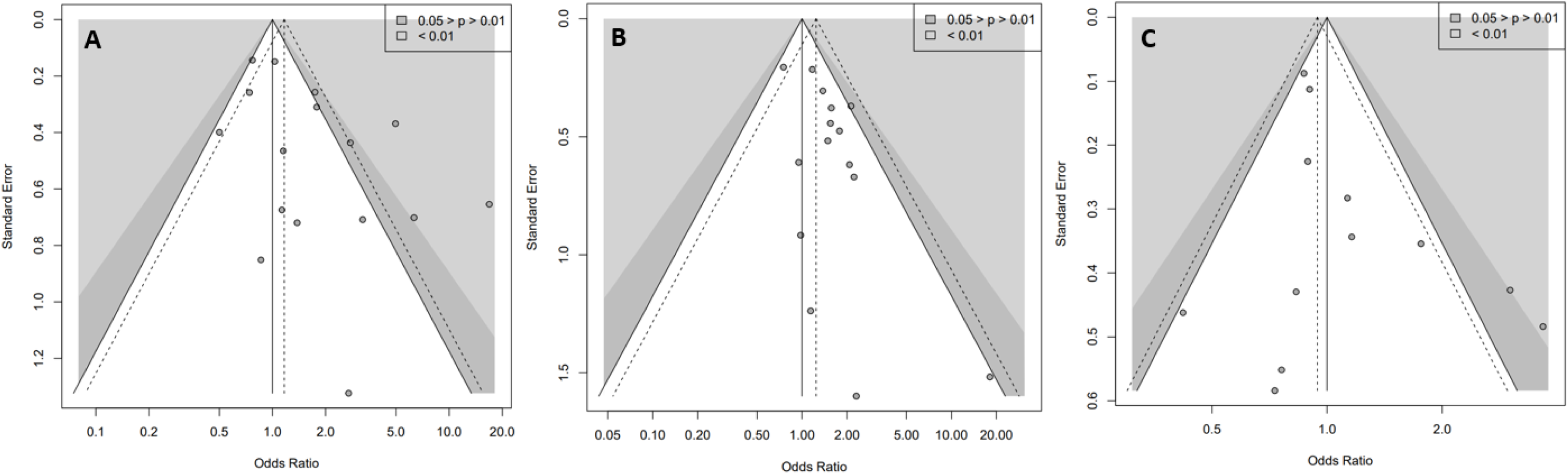
Funnel plots of subgroups by type of statin -. Funnel plot of the association between, A [SLCO1B1 – Simvastatin – SAMS k = 16], B [SLCO1B1 – Atorvastatin – SAMS k = 15], C [SLCO1B1 – Rosuvastatin – SAMS k = 12]. k: number of point estimates. In the funnel plots, each point is an estimation of the associations. The white, dark and light grey zones stand for a p-value of the odds ratio i) non-significant, ii) between 0.05 and 0.01, and iii) <0.01, respectively. The dashed triangle stands for the estimation of the meta-analysis of the association, without adjusting for a potential publication bias.

The sensitivity analysis with RoBMA showed weak evidence of publication bias for simvastatin BF_Publication-bias_ = 2.73 and atorvastatin BF_Publication-bias_ =3.13. We did not perform this analysis for rosuvastatin because of the result of the Egger’s test.

#### Evaluation of the impact of publication bias in secondary analysis by statin type

Only simvastatin and atorvastatin uncorrected meta-analyses were statistically significant (Table 4), with substantial heterogeneity for simvastatin (Cochran’s Q test p=0.02, I^2^=77%) and atorvastatin (Cochran’s Q test p=0.02, I^2^=5%) (S7). Correction for publication bias with the T&F’s method resulted in the loss of association for simvastatin (OR_Trim&Fill_=1.00, CI 95% 0.58–1.72) and atorvastatin (OR_Trim&Fill_=1.13, CI 95% 0.89–1.43). Correcting for publication bias led to a loss of the association for simvastatin and atorvastatin with a 40% and 16% nonsignificant change in the association estimate, respectively (Table 3).

The corrected meta-analysis for simvastatin (OR_RoBMA_=1.11, CI 95% 0.91–2.08) and atorvastatin (OR_RoBMA_=1.02 CI 95% 0.93–1.36) using RoBMA was also inconclusive. Correcting for publication bias led to a loss of the association, with a 34% and 24% nonsignificant change in the association estimate, respectively (Table 3).

#### Detection of the presence of publication bias by SNP type

When subgrouping according to SNP type, we included 42 point estimates for rs4149056, 14 point estimates for rs2306283, and 6 point estimates for rs4363657 with substantial heterogeneity for rs4149056 (Cochran’s Q test p<0.001, I^2^=65%) and moderate heterogeneity for rs2306283 (Cochran’s Q test p=0.5, I^2^=35%) and rs4363657 (Cochran’s Q test p=0.1, I^2^=0%) (S7). Visual inspection of the funnel plot for rs4149056 was suggestive of publication bias (Figure 4), consistent with Egger’s test (Table 2). Conversely, it was not suggestive for SNP rs2306283 with nonsignificant Egger’s test (p=0.8) and undeterminable for rs4363657.

**Figure 4:**
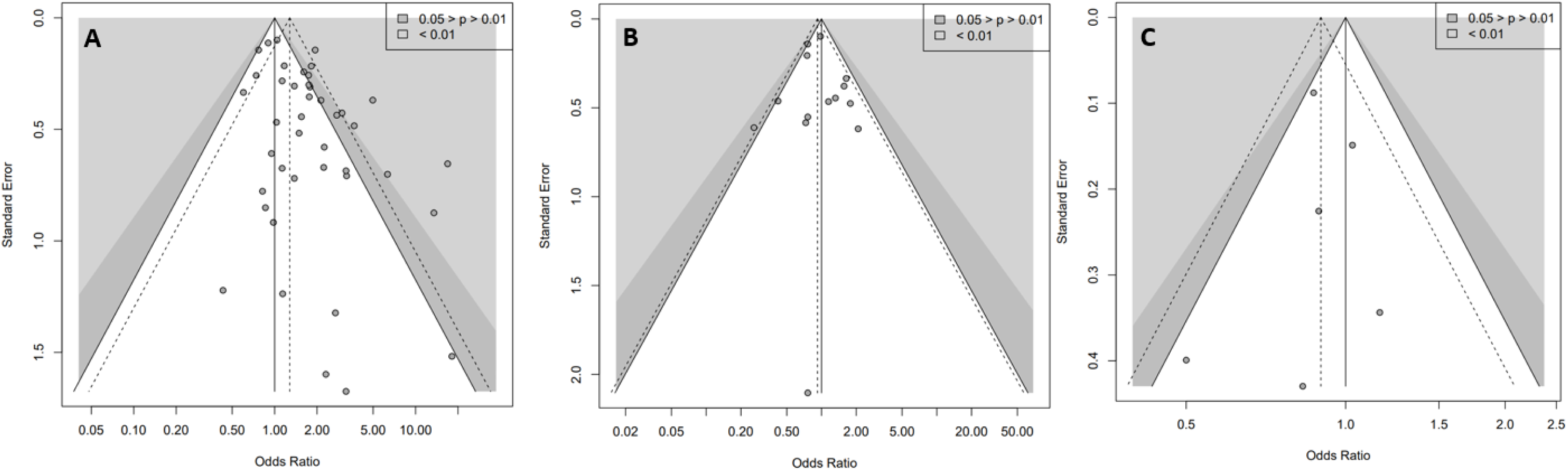
Funnel plots of subgroups by type of SNP -. Funnel plot of the association between A [rs4149056– Statins – SAMS k = 42], B [rs2306283– Statins – SAMS k = 14], C [rS4363657 – Statins – SAMS k = 6]. k: number of point estimates. In the funnel plots, each point is an estimation of the associations. The white, dark and light grey zones stand for a p value of the odds ratio i) non-significant, ii) between 0.05 and 0.01, and iii) <0.01, respectively. The dashed triangle stands for the estimation of the meta-analysis of the association, without adjusting for a potential publication bias.

The sensitivity analysis with RoBMA showed moderate evidence of publication bias (BF_Publication bias_=6) for rs4149056. We did not perform this analysis for SNP rs2306283 and rs4363657.

#### Evaluation of the impact of publication bias by SNP type

In the uncorrected meta-analysis, we observed a significant association only for rs4149056 (OR_Uncorerrected_=1.63, CI 95% 1.34–1.99). Correction for publication bias with the T&F’s method resulted in loss of the association (OR_Trim&Fill_=1.23, CI 95% 0.96–1.58) and a 25% nonsignificant change in the association estimate (ROR_Trim&Fill_= 0.76, CI 95% 0.55–1.04) (Table 3).

The correction using RoBMA also resulted in loss of the significance of the association (OR_RoBMA_=1.11, CI 95% 1.00–1.77) and a 32% significant change in the association estimate (ROR_RoBMA_=0.68, CI 95% 0.55–0.85) (Table 3).

### Other secondary analyses

For secondary analyses focusing on rs4149056, visual inspection of the funnel plots suggested the presence of publication bias for simvastatin, atorvastatin, mixed statin, heterozygous, and homozygous comparisons (S8-9). Egger’s tests were significant only for simvastatin and heterozygous comparisons, and RoBMA was in favor of publication bias. The corrected meta-analysis for simvastatin (OR_Trim&Fill_=1.04, CI 95% 0.55-1.96) and heterozygous comparison (OR_Trim&Fill_=1.04, CI 95% 0.75-1.44) was inconclusive, as was the sensitivity analysis using RoBMA (S10).

### Exploratory analyses

The results of the exploratory analysis do not suggest the presence of any notable publication bias, either in terms of type of statin used or study sponsor and ethnic group (S11–13).

## DISCUSSION

### Summary of evidence and implication

In our meta-epidemiologic study, we demonstrated the presence of publication bias in the pharmacogenetics of SAMS. This publication bias was sufficient to explain the suggested association. Our findings are corroborated by the sensitivity analyses using state of the art methods. To our knowledge, our meta-epidemiologic study is the first to specifically assess publication bias in pharmacogenetics of SAMS.

Previous meta-analyses generally used a small number of estimates (*i.e.*, <10) or did not assess publication bias [36–41]. This may explain why this publication bias was not brought to light earlier. Nevertheless, Jiang *et al.* reported potential publication bias for the heterozygote comparison and the dominant model, but did not use a method to correct this bias [18]. We also detected publication bias for the heterozygote comparison, and correction of this resulted in a loss of the association (Table 2, Table 3). However, we were not able to assess publication bias for the dominant model, due to the low number of estimates (k = 5). Xiang *et al.* highlighted potential publication bias for the heterozygote comparison, the dominant and allelic model, and corrected this bias with the trim and fill method, which resulted in a loss of association for the heterozygote comparison [19]. As for Jiang *et al.*, we found publication bias for the homozygous comparison and the correction also resulted in the loss of the association. However, we did not perform the test for the allelic model due to the low number of associations (k = 8), like the dominant model (k = 5). Their studies may have lacked the power to detect global publication bias in pharmacogenetics of SAMS, but remain consistent with our results.

Consequently, our results cast doubt on the validity of adjusting statin dose or type according to SLCO1B1 genotype in patients in daily clinical practice. The association between SLCO1B1 polymorphisms and SAMS needs to be specifically evaluated in a methodologically appropriate study before further investigating its clinical use, as it may be a false-positive. The pharmacogenetic guidelines should take the publication bias into account when formulating guidance. Furthermore, all pharmacogenetic clinical studies should also be *a priori* registered and systematically published, independent of their results.

### Strengths of the study

In our study, we assessed any publication bias using many point estimates (*i.e.* between 10 and 62), with different levels of heterogeneity (*i.e.* I^2^ between 0 % and 79 %), with complementary methods such as funnel plots, Egger’s test, T&F’s method. To make our publication bias assessment more reliable, we extended our analyses to the RoBMA method. This recent approach has several advantages: first, the Bayesian framework allows us to provide evidence for or against publication bias without having to make all-or-nothing decisions of frequentist publication bias tests with BF [28]. Secondly, this method uses multiple selection models, which are considered the most reliable methods for detecting publication bias when there is significant heterogeneity [30]. RoBMA also provides a more insightful interpretation of results with the use of BF, which provides a continuous measure of evidence for the presence or absence of effect, heterogeneity and publication bias. This approach gave supportive evidence for publication bias, reinforcing our previous findings using conventional techniques. We corrected publication bias using T&F method and RoBMA, which resulted in loss of the association in the main analysis and in the subgroups studied. These results raise questions about the strength of the association originally reported and underscore the critical importance of assessing publication bias in meta-analyses. The observed loss of association may be explained by the fact that some negative or less pronounced estimates in favor of an increased risk of SAMS may have been underrepresented or less frequently published, leading to an overestimation of the association in the literature. Other factors, such as publication practices or P-hacking, may have exacerbated publication bias too [42]. The source of publication bias was also investigated. We examined the asymmetry of the funnel plots showing the main association by statin type or sponsor of the original clinical trials. We observed that simvastatin contributed to the asymmetry of the funnel plot. Analysis by funding type showed that estimates from industry-funded trials had a non-significative trend (S12).

### Limitations

Our study is subject to several limitations. Regarding the limitations of the evidence included in the review, all trials have an observational design for testing the association between a potential biomarker and the treatment effect. Therefore, the potential treatment effect modification is estimated with the corresponding high risk of bias, not protected by randomization. Considering the limits of the review process itself, first, none of the publication bias correction methods can be considered fully reliable, as the actual data generation process remains unknown. Factors, such as the degree of P-hacking, the degree of publication bias and heterogeneity may influence the results. Therefore, despite our efforts to correct publication bias, it is important to remain cautious when interpreting the results and to recognize the inherent limitations of any statistical correction for this type of bias. Secondly, use of many point estimates has resulted in significant heterogeneity, which may have influenced the detection of publication bias using classical symmetry methods – funnel plots, Egger’s test and T&F –, but we obtained consistent results with RoBMA, which strengthens the credibility of our conclusions. Thirdly, we did not use the T&F and RoBMA methods if Egger’s test was not significant. A negative regression test does not guarantee the absence of publication bias [43]. We could have used a 10% significance level to compensate for its low power [24]. This would have made the analyses of several subgroups significant (Table 3, S10), but we preferred to minimize the risk of false positives. In addition, searching for interactions by ROR calculation is associated with a lack of power [44]. Nevertheless, the results of our study are all the stronger for having underestimated publication bias in most of our analyses. In addition, the effect of publication bias was more pronounced for simvastatin, which is known to be a more important substrate of the OATP transporter (SLCO1B1). It would have been interesting to also evaluate the BCRP (ABCG2) transporter with other statins less affected by publication bias that are also substrates of this transporter. This could be an important addition to a future study. We chose to focus on SCLO1B1 because it is the focus of the recent CPIC recommendations.

## Conclusion

In this meta-epidemiological study, we provide a cluster of consistent evidence of publication bias in the SLCO1B1- SAMS association. Our analysis revealed the presence of publication bias, which after correction, resulted in the loss of significance of the association. Given the limitations of publication bias adjustment methods, these results do not definitively rule out this association, but they call it into question. This publication bias should be considered in further research. Indeed, failing to consider the risk of publication bias in this example might lead to failure to treat patients who stand to benefit from statins.

## Data Availability

The protocol was registered a priori on the Open Science Framework

https://osf.io/36jq8/

## Acknowledgment

We would like to thank Gaëlle Siméon for her help in reviewing this article.

## Author Contributions

Conceptualization

AG, GG, FG, AL, IA. Data curation: AG, IA, SG. Formal analysis: AG Investigation: AG, GG, FG, AL, JCL. Methodology: AG, GG, FG, AL. Writing– original draft: AG, GG. Writing– review & editing: AG, GG, FG, AL, IA, SG, JCL, JMW, CV

## Non-standard Abbreviations and Acronyms

LDL-C: Low-density lipoprotein cholesterol
RCT: Randomized controlled trial
CV: Cardiovascular
AE: Adverse event
SAMS: Statin-associated muscle symptoms
CK: Creatine kinase
SLCO1B1: Solute carrier organic anion transporter family member 1B1
EMA: European Medicines Agency
FDA: Food and Drug Administration
CPIC: Clinical Pharmacogenetics Implementation Consortium
T&F: Trim-and-Fill
NOS: Newcastle-Ottawa scale
SNP: Single nucleotide polymorphism
PRISMA: Preferred Reporting Items for Systematic Reviews and Meta-Analyses
OR: Odds ratio
CI: Confidence interval
OR_Trim&Fill_: OR corrected for publication bias by Trim and Fill method
ROR_Trim&Fill_: Ratio of odds ratio ( OR corrected by Trim and Fill method)
OR_Uncorrected_: OR uncorrected for publication bias
RoBMA: Robust Bayesian Meta-Analysis
BF: Bayes factor
OR_RoBMA_: OR corrected for publication bias by RoBMA approach
ROR_RoBMA_: Ratio of odds ratio ( OR corrected by RoBMA approach)
SNP: Single Nucleotide Polymorphism
WT: Wild type

## Supplemental Material

### Supplementary material 1 (S1): PRISMA check list

| Section and Topic | Item # | Checklist item | Location where item is reported |
| --- | --- | --- | --- |
| **TITLE** |  |  |  |
| Title | 1 | Identify the report as a systematic review. | Title |
| **ABSTRACT** |  |  |  |
| Abstract | 2 | See the PRISMA 2020 for Abstracts checklist. | Abstract (348 words) |
| **INTRODUCTION** |  |  |  |
| Rationale | 3 | Describe the rationale for the review in the context of existing knowledge. | INTRODUCTION |
| Objectives | 4 | Provide an explicit statement of the objective(s) or question(s) the review addresses. | INTRODUCTION |
| **METHODS** |  |  |  |
| Eligibility criteria | 5 | Specify the inclusion and exclusion criteria for the review and how studies were grouped for the syntheses. | Search strategy and inclusion criteria |
| Information sources | 6 | Specify all databases, registers, websites, organisations, reference lists and other sources searched or consulted to identify studies. Specify the date when each source was last searched or consulted. | Search strategy and inclusion criteria |
| Search strategy | 7 | Present the full search strategies for all databases, registers and websites, including any filters and limits used. | Supplementary 1 (S1) : Search strategy |
| Selection process | 8 | Specify the methods used to decide whether a study met the inclusion criteria of the review, including how many reviewers screened each record and each report retrieved, whether they worked independently, and if applicable, details of automation tools used in the process. | Search strategy and inclusion criteria |
| Data collection process | 9 | Specify the methods used to collect data from reports, including how many reviewers collected data from each report, whether they worked independently, any processes for obtaining or confirming data from study investigators, and if applicable, details of automation tools used in the process. | Data extraction and quality assessment of included studies |
| Data items | 10a | List and define all outcomes for which data were sought. Specify whether all results that were compatible with each outcome domain in each study were sought (e.g. for all measures, time points, analyses), and if not, the methods used to decide which results to collect. | Data extraction and quality assessment of included studies |
|  | 10b | List and define all other variables for which data were sought (e.g. participant and intervention characteristics, funding sources). Describe any assumptions made about any missing or unclear information. | Data extraction and quality |
|  |  |  | assessment of included studies |
| Study risk of bias assessment | 11 | Specify the methods used to assess risk of bias in the included studies, including details of the tool(s) used, how many reviewers assessed each study and whether they worked independently, and if applicable, details of automation tools used in the process. | Data extraction and quality assessment of included studies |
| Effect measures | 12 | Specify for each outcome the effect measure(s) (e.g. risk ratio, mean difference) used in the synthesis or presentation of results. | Dataset |
| Synthesis methods | 13a | Describe the processes used to decide which studies were eligible for each synthesis (e.g. tabulating the study intervention characteristics and comparing against the planned groups for each synthesis (item #5)). | Outcome |
|  | 13b | Describe any methods required to prepare the data for presentation or synthesis, such as handling of missing summary statistics, or data conversions. | Statistical analysis |
|  | 13c | Describe any methods used to tabulate or visually display results of individual studies and syntheses. | Statistical analysis |
|  | 13d | Describe any methods used to synthesize results and provide a rationale for the choice(s). If meta-analysis was performed, describe the model(s), method(s) to identify the presence and extent of statistical heterogeneity, and software package(s) used. | Statistical analysis |
|  | 13e | Describe any methods used to explore possible causes of heterogeneity among study results (e.g. subgroup analysis, meta-regression). | Statistical analysis |
|  | 13f | Describe any sensitivity analyses conducted to assess robustness of the synthesized results. | Statistical analysis |
| Reporting bias assessment | 14 | Describe any methods used to assess risk of bias due to missing results in a synthesis (arising from reporting biases). | Statistical analysis |
| Certainty assessment | 15 | Describe any methods used to assess certainty (or confidence) in the body of evidence for an outcome. | Statistical analysis |
| **RESULTS** |  |  |  |
| Study selection | 16a | Describe the results of the search and selection process, from the number of records identified in the search to the number of studies included in the review, ideally using a flow diagram. | Figures: flow chart |
|  | 16b | Cite studies that might appear to meet the inclusion criteria, but which were excluded, and explain why they were excluded. | Figures: flow chart |
| Study characteristics | 17 | Cite each included study and present its characteristics. | Tables: characteristics of include studies |
| Risk of bias in studies | 18 | Present assessments of risk of bias for each included study. | Supplementary 5.(1-2): Risk of bias assessment in selected |
|  |  |  | systematic review |
| Results of individual studies | 19 | For all outcomes, present, for each study: (a) summary statistics for each group (where appropriate) and (b) an effect estimate and its precision (e.g. confidence/credible interval), ideally using structured tables or plots. | X |
| Results of syntheses | 20a | For each synthesis, briefly summarise the characteristics and risk of bias among contributing studies. | Supplementary 5.(1-2): Risk of bias assessment in selected systematic review |
|  | 20b | Present results of all statistical syntheses conducted. If meta-analysis was done, present for each the summary estimate and its precision (e.g. confidence/credible interval) and measures of statistical heterogeneity. If comparing groups, describe the direction of the effect. | Tables: results |
|  | 20c | Present results of all investigations of possible causes of heterogeneity among study results. | Assessment of publication bias in the main association<br>Assessment of publication bias in subgroup |
|  | 20d | Present results of all sensitivity analyses conducted to assess the robustness of the synthesized results. | Assessment of publication bias in the main association<br>Assessment of publication bias in subgroup |
| Reporting biases | 21 | Present assessments of risk of bias due to missing results (arising from reporting biases) for each synthesis assessed. | Assessment of publication bias in the main association<br>Assessment of publication bias in subgroup |
| Certainty of | 22 | Present assessments of certainty (or confidence) in the body of evidence for each outcome assessed. | Assessment of |
| evidence |  |  | publication bias in the main association<br>Assessment of publication bias in subgroup |
| **DISCUSSION** |  |  |  |
| Discussion | 23a | Provide a general interpretation of the results in the context of other evidence. | Summary of evidence Strengths of the study |
|  | 23b | Discuss any limitations of the evidence included in the review. | Limitations |
|  | 23c | Discuss any limitations of the review processes used. | Limitations |
|  | 23d | Discuss implications of the results for practice, policy, and future research. | Summary of evidence |
| **OTHER INFORMATION** |  |  |  |
| Registration and protocol | 24a | Provide registration information for the review, including register name and registration number, or state that the review was not registered. | Protocol and registration |
|  | 24b | Indicate where the review protocol can be accessed, or state that a protocol was not prepared. | Protocol and registration |
|  | 24c | Describe and explain any amendments to information provided at registration or in the protocol. | Protocol and registration |
| Support | 25 | Describe sources of financial or non-financial support for the review, and the role of the funders or sponsors in the review. | Sources of Funding |
| Competing interests | 26 | Declare any competing interests of review authors. | Disclosures |
| Availability of data, code and other materials | 27 | Report which of the following are publicly available and where they can be found: template data collection forms; data extracted from included studies; data used for all analyses; analytic code; any other materials used in the review. | Supplemental Material |

From: Page MJ, McKenzie JE, Bossuyt PM, Boutron I, Hoffmann TC, Mulrow CD, et al. The PRISMA 2020 statement: an updated guideline for reporting systematic reviews. BMJ 2021;372:n71. doi: 10.1136/bmj.n71

### Supplementary material (S2): Search strategy

*<u>Pharmacogenomics keywords</u>* : (”polymorphism, genetic”[MeSH Terms] OR (”polymorphism”[All Fields] AND “genetic”[All Fields]) OR “genetic polymorphism”[All Fields] OR (”polymorphism”[All Fields] AND “genetic”[All Fields]) OR “polymorphism genetic”[All Fields] OR ((”organic anion transporters”[MeSH Terms] OR (”organic”[All Fields] AND “anion”[All Fields] AND “transporters”[All Fields]) OR “organic anion transporters”[All Fields] OR (”solute”[All Fields] AND “carrier”[All Fields] AND “organic”[All Fields] AND “anion”[All Fields]) OR “solute carrier organic anion”[All Fields]) AND (”biological transport”[MeSH Terms] OR (”biological”[All Fields] AND “transport”[All Fields]) OR “biological transport”[All Fields] OR “transport”[All Fields] OR “membrane transport proteins”[MeSH Terms] OR (”membrane”[All Fields] AND “transport”[All Fields] AND “proteins”[All Fields]) OR “membrane transport proteins”[All Fields] OR “transporter”[All Fields] OR “transporters”[All Fields] OR “transportable”[All Fields] OR “transportation”[MeSH Terms] OR “transportation”[All Fields] OR “transportations”[All Fields] OR “transported”[All Fields] OR “transporter s”[All Fields] OR “transporting”[All Fields] OR “transports”[All Fields]) AND “1B1”[All Fields]) OR “SLCO1B1”[All Fields] OR ((”organic anion transporters”[MeSH Terms] OR (”organic”[All Fields] AND “anion”[All Fields] AND “transporters”[All Fields]) OR “organic anion transporters”[All Fields] OR (”organic”[All Fields] AND “anion”[All Fields] AND “transporter”[All Fields]) OR “organic anion transporter”[All Fields]) AND (”peptides”[MeSH Terms] OR “peptides”[All Fields] OR “polypeptide”[All Fields] OR “polypeptides”[All Fields] OR “polypeptid”[All Fields] OR “polypeptide s”[All Fields] OR “polypeptidic”[All Fields]) AND “1B1”[All Fields]) OR “OATP1B1”[All Fields])

AND

*<u>Statin keywords</u>* : statin OR rosuvastatin OR fluvastatin OR pravastatin (”hydroxymethylglutaryl coa reductase inhibitors”[Pharmacological Action] OR “hydroxymethylglutaryl coa reductase inhibitors”[MeSH Terms] OR (”hydroxymethylglutaryl coa”[All Fields] AND “reductase”[All Fields] AND “inhibitors”[All Fields]) OR “hydroxymethylglutaryl coa reductase inhibitors”[All Fields] OR “statin”[All Fields] OR “statins”[All Fields] OR “statin s”[All Fields] OR “statine”[Supplementary Concept] OR “statine”[All Fields] OR “statines”[All Fields] OR (”rosuvastatin calcium”[MeSH Terms] OR (”rosuvastatin”[All Fields] AND “calcium”[All Fields]) OR “rosuvastatin calcium”[All Fields] OR “rosuvastatin”[All Fields]) OR (”fluvastatin”[MeSH Terms] OR “fluvastatin”[All Fields]) OR (”pravastatin”[MeSH Terms] OR “pravastatin”[All Fields] OR “pravastatin s”[All Fields]) OR (”simvastatin”[MeSH Terms] OR “simvastatin”[All Fields] OR “simvastatin s”[All Fields] OR “simvastatins”[All Fields]) OR (”cerivastatin”[Supplementary Concept] OR “cerivastatin”[All Fields] OR “cerivastatin s”[All Fields]) OR (”lovastatin”[MeSH Terms] OR “lovastatin”[All Fields] OR “lovastatine”[All Fields] OR “lovastatin s”[All Fields]) OR (”atorvastatin”[MeSH Terms] OR “atorvastatin”[All Fields] OR “atorvastatine”[All Fields] OR “atorvastatin s”[All Fields]) OR (”pitavastatin”[Supplementary Concept] OR “pitavastatin”[All Fields]))

AND

*<u>Adverse event keywords</u>* : (”muscular diseases”[MeSH Terms] OR (”muscular”[All Fields] AND “diseases”[All Fields]) OR “muscular diseases”[All Fields] OR “myopathies”[All Fields] OR “myopathy”[All Fields] OR (”myalgia”[MeSH Terms] OR “myalgia”[All Fields] OR “myalgias”[All Fields]) OR (”myositis”[MeSH Terms] OR “myositis”[All Fields] OR “myositides”[All Fields]) OR (”rhabdomyolysis”[MeSH Terms] OR “rhabdomyolysis”[All Fields] OR “rhabdomyolyses”[All Fields]) OR (”creatine kinase”[MeSH Terms] OR (”creatine”[All Fields] AND “kinase”[All Fields]) OR “creatine kinase”[All Fields]) OR “CK”[All Fields])

#### Search strategy PubMed details

*<u>Pharmacogenomics keywords</u>* : (’polymorphism genetic’/exp OR ’polymorphism genetic’ OR ((’polymorphism’/exp OR polymorphism) AND (’genetic’/exp OR genetic)) OR ’solute carrier organic anion transporter 1b1’/exp OR ’solute carrier organic anion transporter 1b1’ OR ((’solute’/exp OR solute) AND (’carrier’/exp OR carrier) AND organic AND (’anion’/exp OR anion) AND (’transporter’/exp OR transporter) AND 1b1) OR slco1b1 OR ’organic anion transporter polypeptide 1b1’ OR (organic AND (’anion’/exp OR anion) AND (’transporter’/exp OR transporter) AND (’polypeptide’/exp OR polypeptide) AND 1b1) OR oatp1b1)

AND

*<u>Statin keywords</u>*: (’statin’/exp OR statin OR ’rosuvastatin’/exp OR rosuvastatin OR ’fluvastatin’/exp OR fluvastatin OR ’pravastatin’/exp OR pravastatin OR ’simvastatin’/exp OR simvastatin OR ’cerivastatin’/exp OR cerivastatin OR ’lovastatin’/exp OR lovastatin OR ’atorvastatin’/exp OR atorvastatin OR ’pitavastatin’/exp OR pitavastatin)

AND

*<u>Adverse event keywords</u>*: (’myopathy’/exp OR myopathy OR ’myalgia’/exp OR myalgia OR ’myositis’/exp OR myositis OR ’rhabdomyolysis’/exp OR rhabdomyolysis OR ’creatine kinase’/exp OR ’creatine kinase’ OR ((’creatine’/exp OR creatine) AND (’kinase’/exp OR kinase)) OR ck)

#### Search strategy Embase details

ID Search Hits

#1 polymorphism genetic

#2 solute carrier organic anion transporter 1B1 #3 SLCO1B1

#4 organic anion transporter polypeptide 1B1 OR OATP1B1 #5 #1 OR #2 OR #3 OR #4

#6 statin

#7 rosuvastatin

#8 pravastatin

#9 simvastatin

#10 cerivastatin

#11 lovastatin

#12 atorvastatin

#13 pitavastatin

#14 #6 OR #7 OR #8 OR #9 OR #10 OR #11 OR #12 OR #13

#15 myopathy

#16 myalgia

#17 myositis

#18 rhabdomyolysis

#19 creatine kinase

#20 CK

#21 #15 OR #16 OR #17 OR #18 OR #19 OR #20

#22 #5 AND #14 #21

#### Search strategy Cochrane Central details

### Supplementary material 3 (S3): Overlap primary studies in systematic reviews and risk of bias assessment in selected systematic review using the Newcastle-Ottawa scale (NOS)

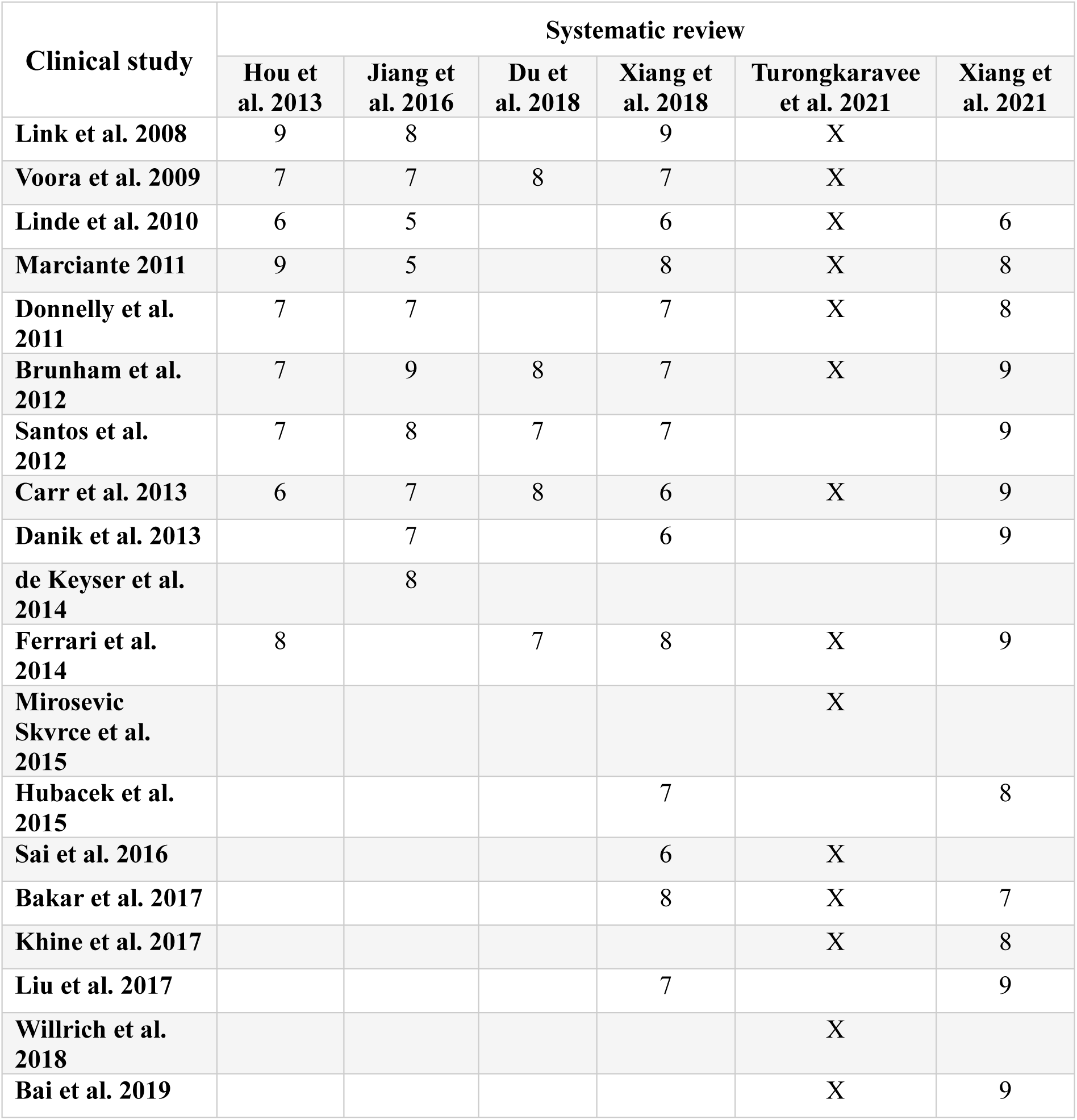

Supplementary material 4 (S4): Formula of the ratio of odds ratio and its 95% confidence interval

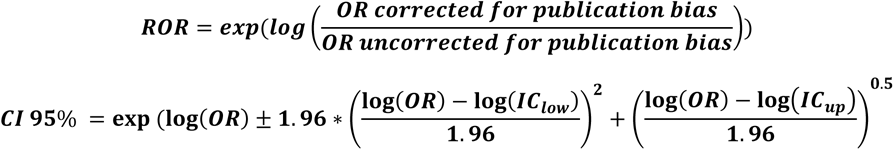

### Supplementary 5 (S5): Assessment of the risk of bias (Score for Genetic Association Studies) in original clinical studies included by Turongkaravee et al. 2021

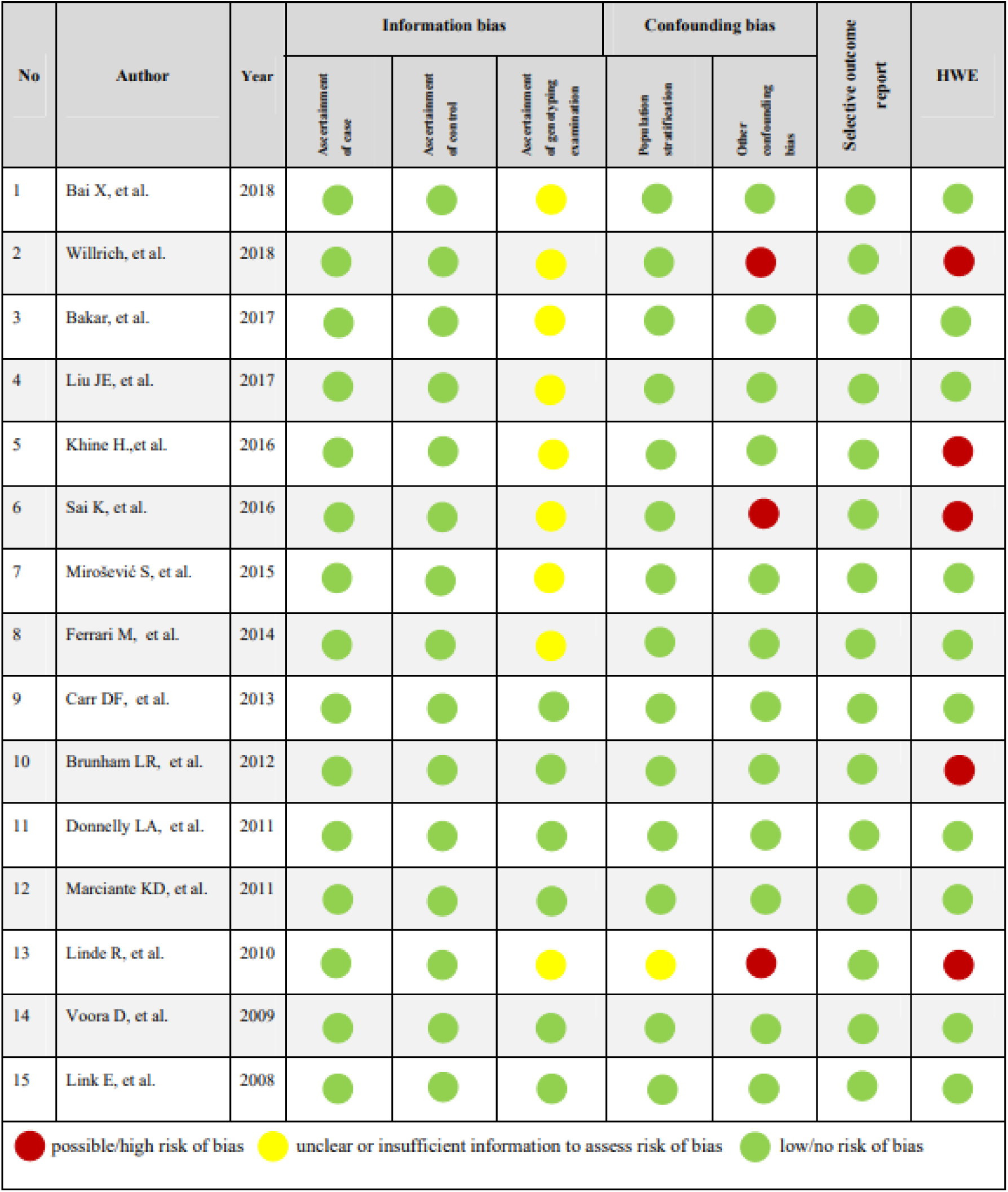

### Supplementary material (S6): Bayes factor for the overall association and subgroups

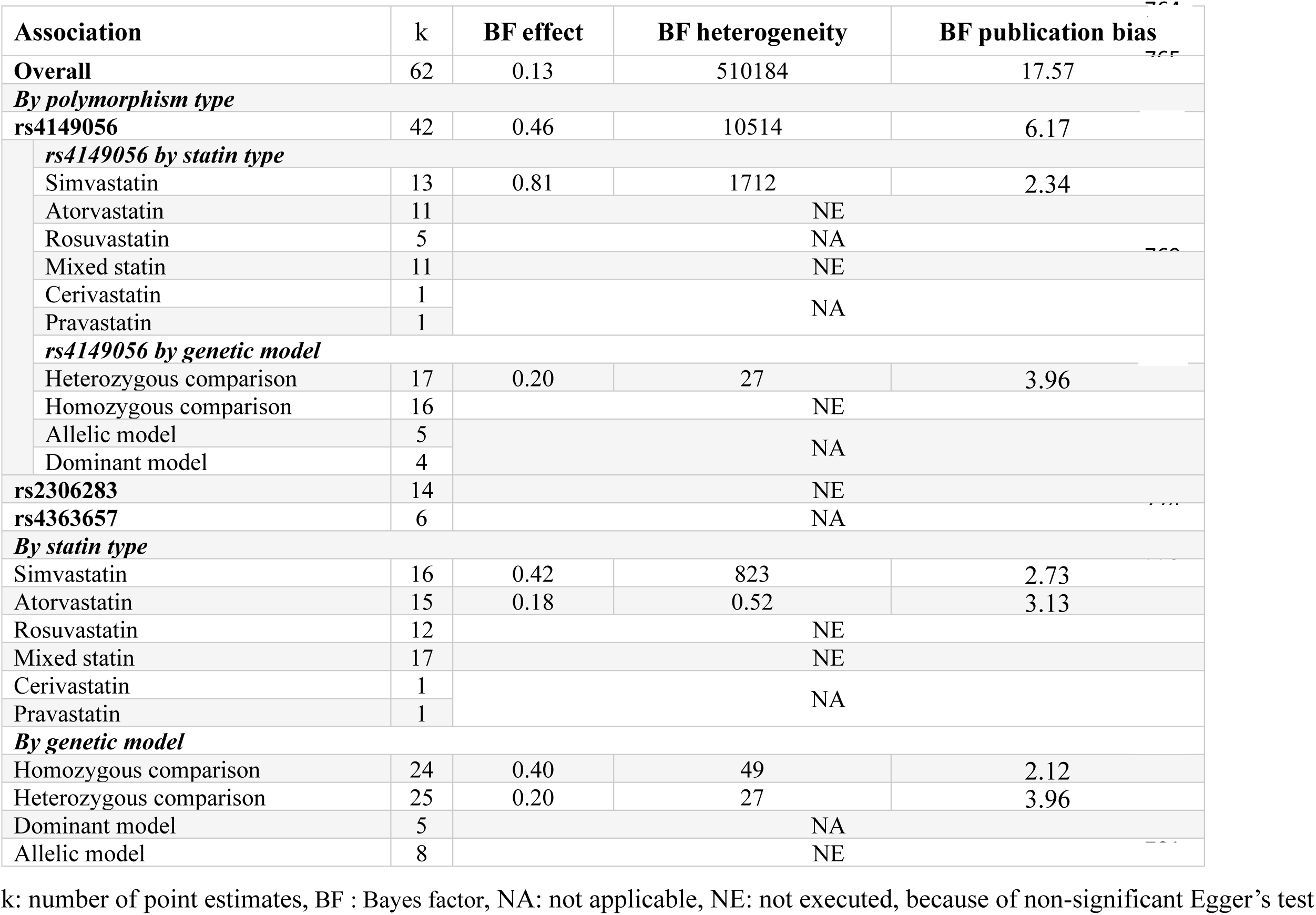

### Supplementary material 7 (S7): Proportion of heterogeneity in primary analysis and secondary analysis

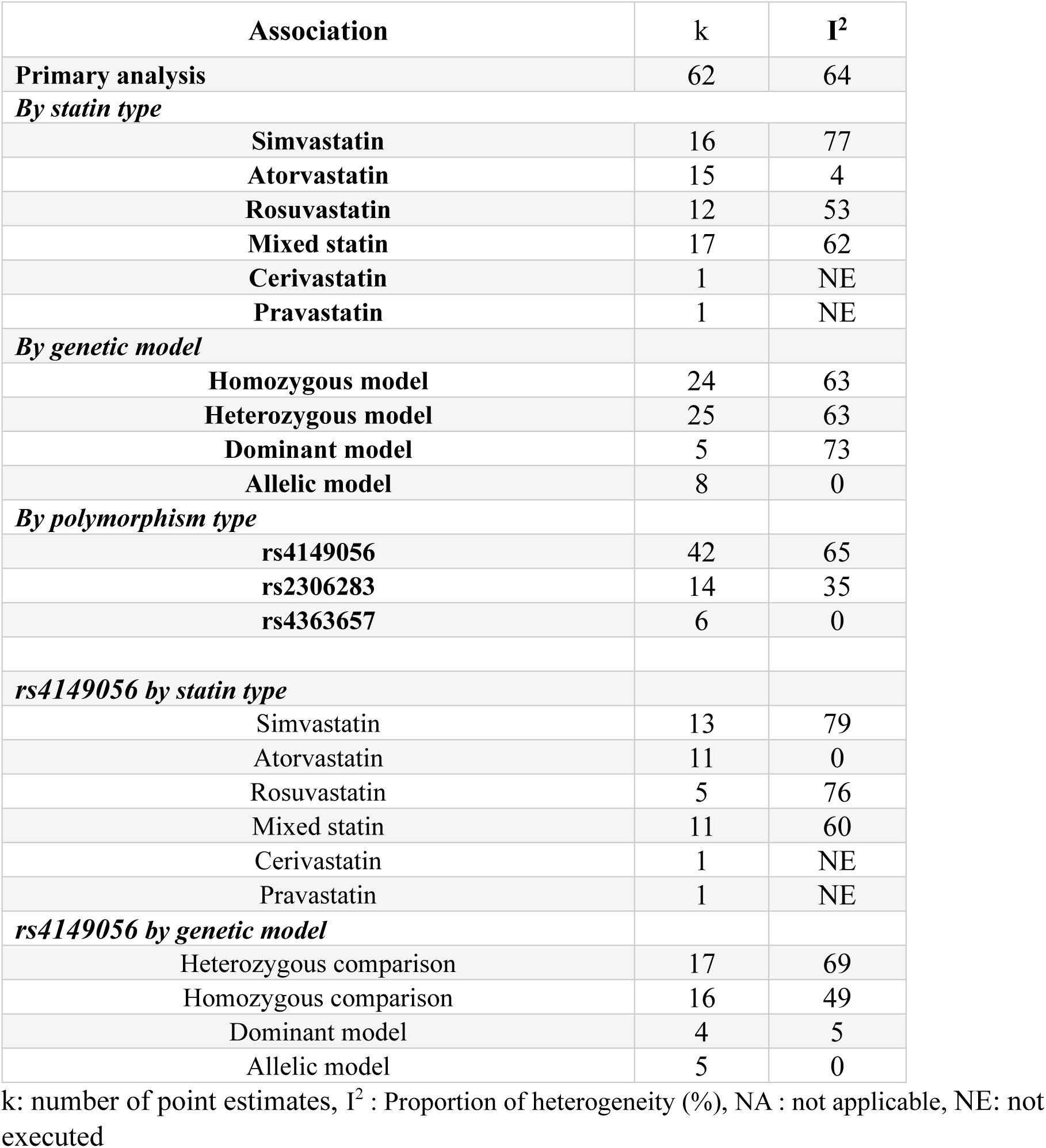

| **Association** | **k** | **I<sup>2</sup>** |
| --- | --- | --- |
| **Primary analysis** | **62** | **64** |
| ***By statin type*** |  |  |
| **Simvastatin** | **16** | **77** |
| **Atorvastatin** | **15** | **4** |
| **Rosuvastatin** | **12** | **53** |
| **Mixed statin** | **17** | **62** |
| **Cerivastatin** | **1** | **NE** |
| **Pravastatin** | **1** | **NE** |
| ***By genetic model*** |  |  |
| **Homozygous model** | **24** | **63** |
| **Heterozygous model** | **25** | **63** |
| **Dominant model** | **5** | **73** |
| **Allelic model** | **8** | **0** |
| ***By polymorphism type*** |  |  |
| **rs4149056** | **42** | **65** |
| **rs2306283** | **14** | **35** |
| **rs4363657** | **6** | **0** |
| ***rs4149056 by statin type*** |  |  |
| **Simvastatin** | **13** | **79** |
| **Atorvastatin** | **11** | **0** |
| **Rosuvastatin** | **5** | **76** |
| **Mixed statin** | **11** | **60** |
| **Cerivastatin** | **1** | **NE** |
| **Pravastatin** | **1** | **NE** |
| ***rs4149056 by genetic model*** |  |  |
| **Heterozygous comparison** | **17** | **69** |
| **Homozygous comparison** | **16** | **49** |
| **Dominant model** | **4** | **5** |
| **Allelic model** | **5** | **0** |

k: number of point estimates, I<sup>2</sup> : Proportion of heterogeneity (%), NA : not applicable, NE: not executed

### Supplementary 8 (S8): Funnel plots of rs4149056 by type of statin

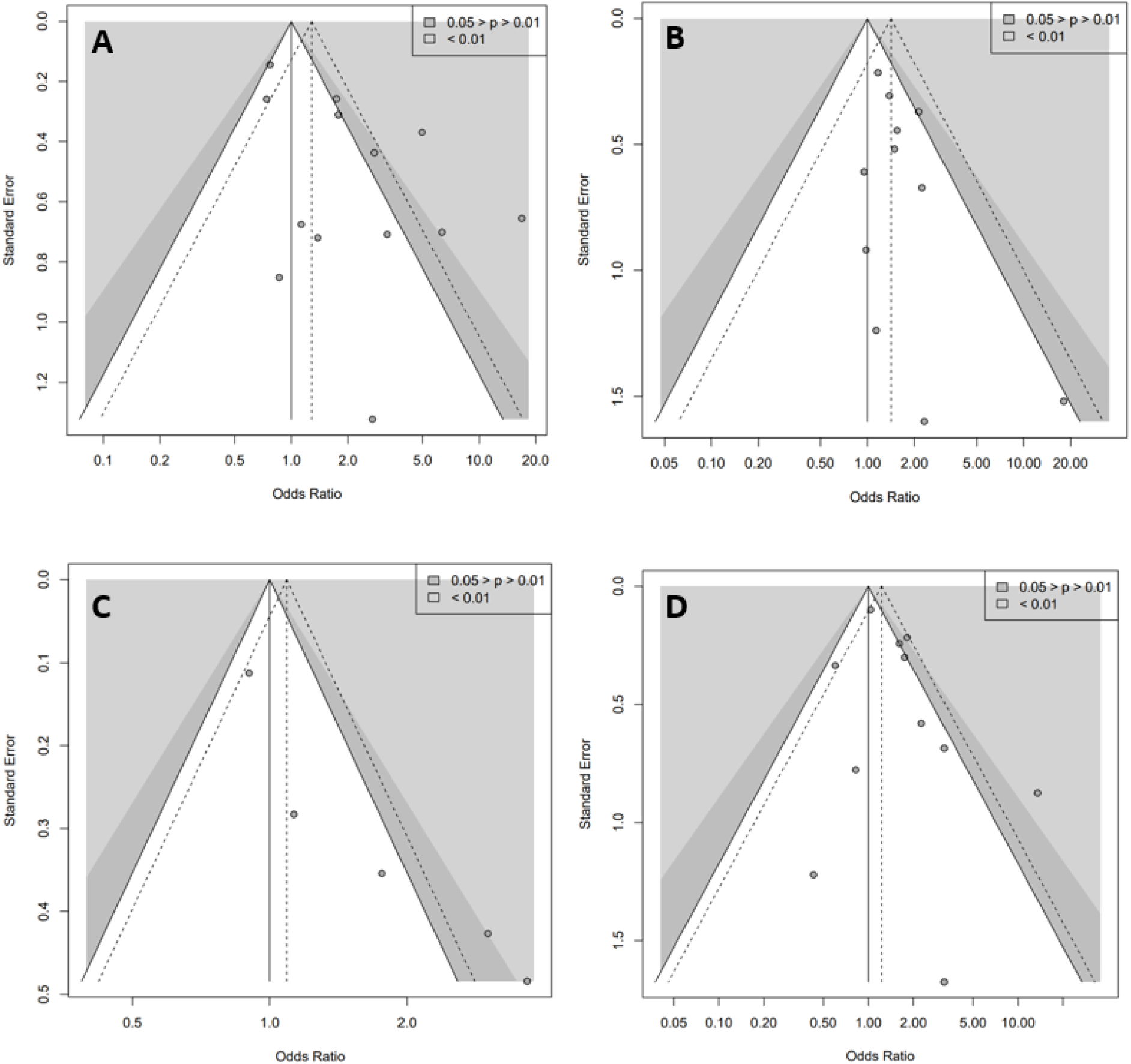
Funnel plots of the association between, A [rs4149056– Simvastatin – SAMS k = 13], B [rs4149056– Atorvastatin – SAMS k = 11], C [rs4149056– Rosuvastatin – SAMS k = 5], D [rs4149056– mixed statin – SAMS k = 11]. k: number of point estimates. In the funnel plots each point is an estimation of the associations. The white, dark and light grey zones stand for a p value of the odds ratio i) non-significant, ii) between 0.05 and 0.01, and iii) <0.01, respectively. The dashed triangle stands for the estimation of the meta-analysis of the association, without adjusting for a potential publication bias.

### Supplementary 9 (S9): Funnel plot of rs4149056 by type of genetic model

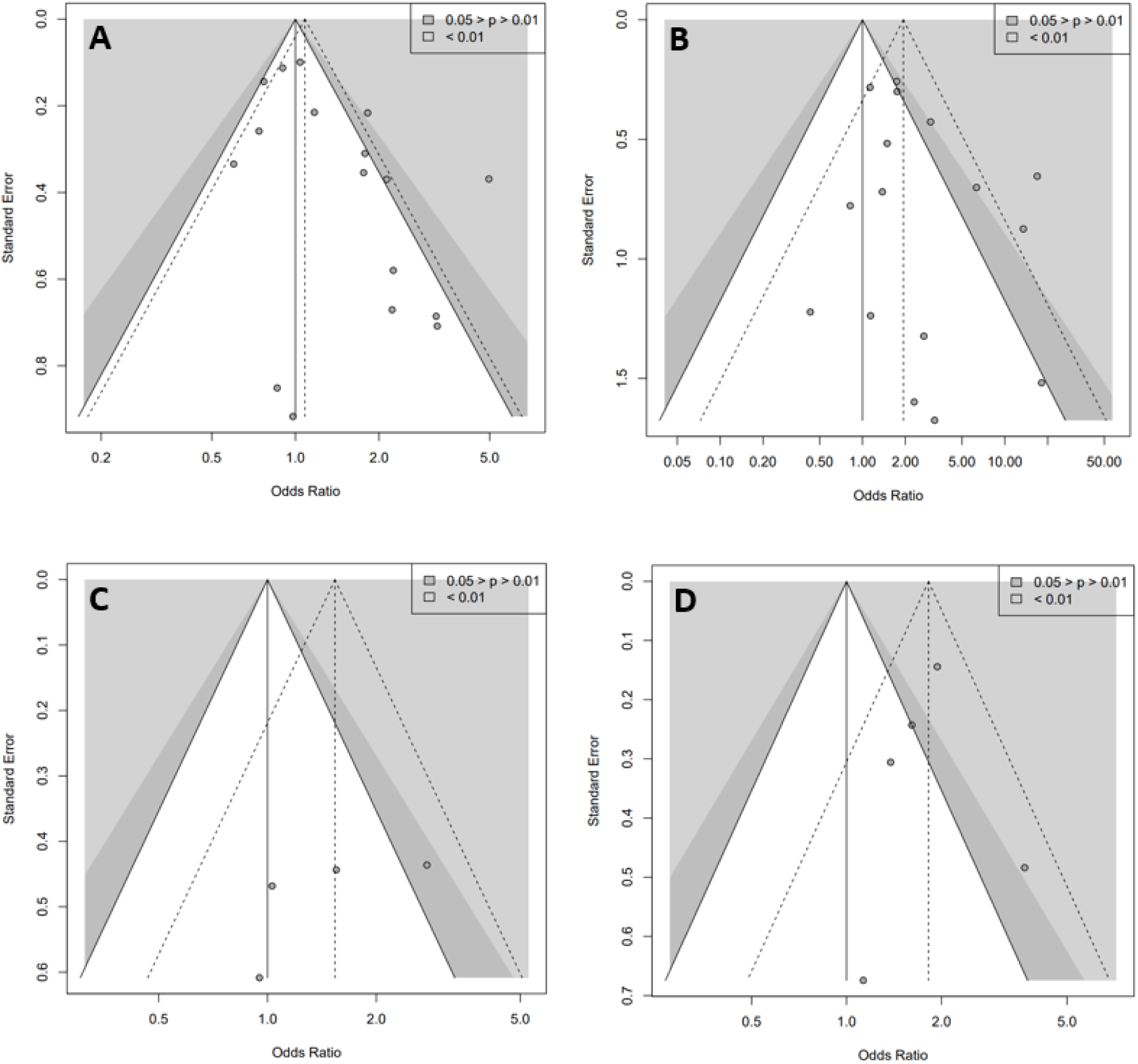
Funnel plot of the association between, A [rs4149056 Heterozygous comparison – Statin – SAMS k = 17], B [rs4149056 Homozygous comparison – Statin – SAMS k = 16], C [rs4149056 Dominant model – Statin – SAMS k = 5], D [rs4149056 Allelic model – Statin – SAMS k = 5]. k: number of point estimates. In the funnel plots each point is an estimation of the associations. The white, dark and light grey zones stand for a p value of the odds ratio i) non-significant, ii) between 0.05 and 0.01, and iii) <0.01, respectively. The dashed triangle stands for the estimation of the meta-analysis of the association, without adjusting for a potential publication bias.

### Supplementary material 10 (S10) : Table of supplementary results for secondary analysis focused on rs4149056

| Association | k | Evaluation of publication bias |  |  | OR<sub>Uncorrected</sub><br>CI95% | OR<sub>Corrected</sub> CI95% |  | Effect of publication bias |  |
| --- | --- | --- | --- | --- | --- | --- | --- | --- | --- |
|  |  | Funnel plot | Egger | RoBMA |  | OR<sub>Trim&Fill</sub> | OR<sub>RoBMA</sub> | ROR<sub>T&F</sub> 95%CI | ROR<sub>RoBMA</sub> 95%CI |
| rs4149056 | 42 | suggestive of a publication bias | 0.001 * | In favor + | 1.63 [1.34 - 1.99] | 1.23 [0.96 - 1.58] | 1.11 [1.00 - 1.77] | 0.76 [0.55 - 1.04] | 0.68 [0.55 - 0.86] |
| By statin type |  |  |  |  |  |  |  |  |  |
| Simvastatin | 13 | suggestive of a publication bias | 0,02 * | In favor | 2.05 [1.24 - 3.41] | 1.04 [0.55 - 1.96] | 1.27 [0.98 - 2.75] | 0.50 [0.22 - 1.14] | 0.62 [0.35 - 1.10] |
| Atorvastatin | 11 | suggestive of a publication bias | 0,1 | NE | 1.41 [1.09 - 1.84] | NE |  |  |  |
| Rosuvastatin | 5 | undeterminable | NA | NE | 1.62 [0.96 - 2.76] |  |  |  |  |
| Mixed statin | 11 | suggestive of a publication bias | 0,1 | NE | 1.47 [1.03 - 2.11] |  |  |  |  |
| Cerivastatin | 1 | NA |  |  |  |  |  |  |  |
| Pravastatin | 1 |  |  |  |  |  |  |  |  |
| By genetic model |  |  |  |  |  |  |  |  |  |
| Heterozygous comparison | 17 | suggestive of a publication bias | 0,02 * | In favor + | 1.37 [1.03 - 1.83] | 1.04 [0.75 - 1.44] | 1.03 [0.94 - 1.49] | 0.75 [0.49 - 1.17] | 0.75 [0.56 - 1.01] |
| Homozygous comparison | 16 | suggestive of a publication bias | 0,1 | NE | 2.35 [1.46 - 3.76] | NE |  |  |  |
| Dominant model | 4 | undeterminable | NA | NE | 1.53 [0.93 - 2.50] |  |  |  |  |
| Allelic model | 5 | undeterminable | NA | NE | 1.82 [1.47 - 2.26] |  |  |  |  |

k: number of point estimates, \* indicates significant p-value, NA: not applicable, NE: not executed, because of non-significant Egger's test, +++ indicates Bayes factor <10, ++ indicates Bayes factor 3-10, + indicates Bayes factor <3, ROR<sub>Trim&Fill</sub>: ratio of OR<sub>Trim&Fill</sub> and OR<sub>Uncorrected</sub>; ROR<sub>RoBMA</sub>: ratio of OR<sub>Robust Bayesian Meta-Analysis</sub> and OR<sub>Uncorrected</sub>

### Supplementary material 11 (S11): Funnel plot of the primary analysis [SLCO1B1 SNPs – Statins – SAMS] based on statin type

**Figure S11:**
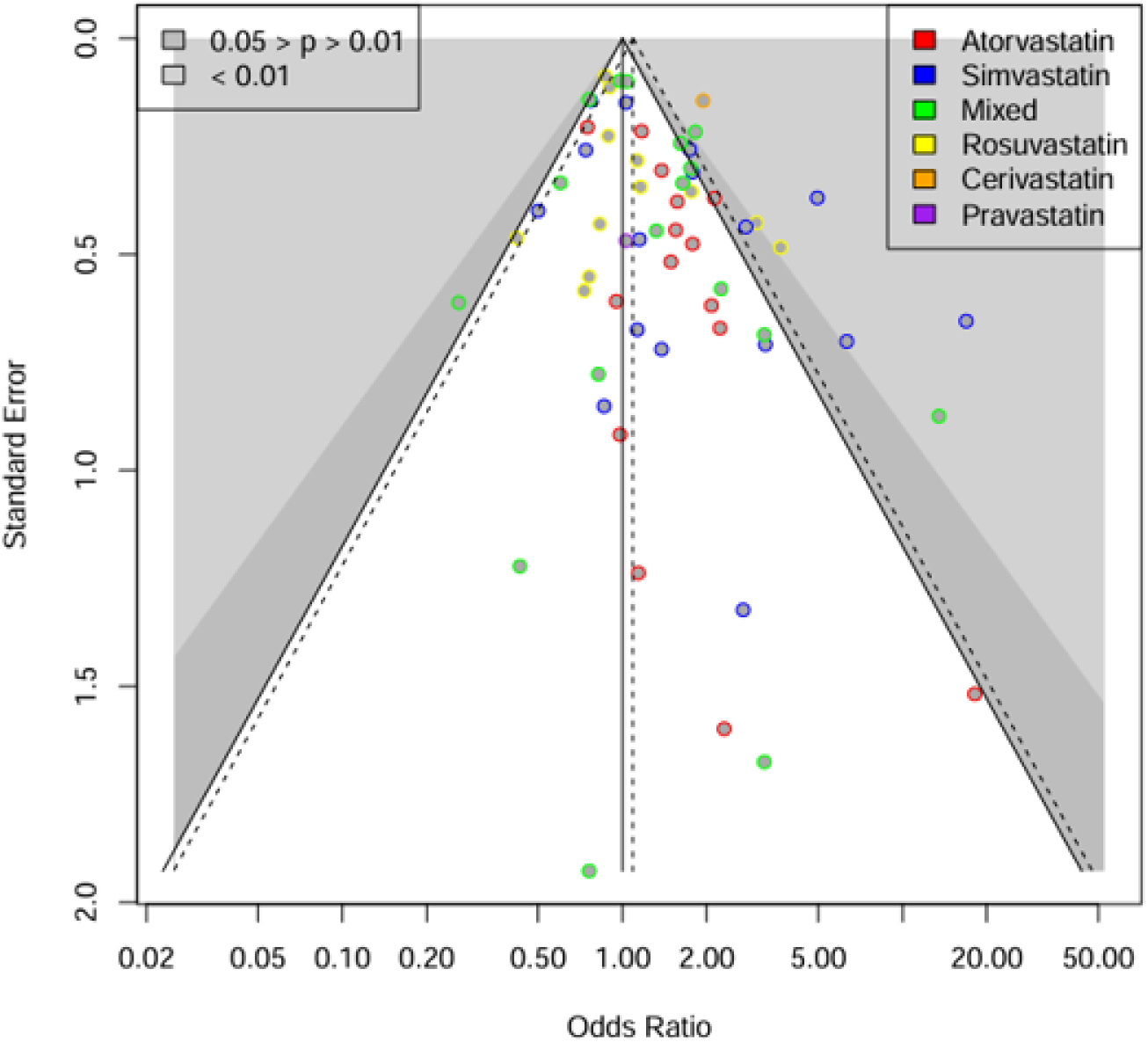
Funnel plot of the primary analysis [SLCO1B1 SNPs – Statins – SAMS k = 62] based on statin type-. k: number of point estimates. In the funnel plots each point is an estimation of the associations, color corresponds to statin type. The white, dark and light grey zones stand for a p value of the odds ratio i) non-significant, ii) between 0.05 and 0.01, and iii) <0.01, respectively. The dashed triangle stands for the estimation of the meta-analysis of the association, without adjusting for a potential publication bias.

### Supplementary material 12 (S12): Funnel plot of the association [SLCO1B1 SNPs – Statins – SAMS] based on study funding

**Figure S12.**
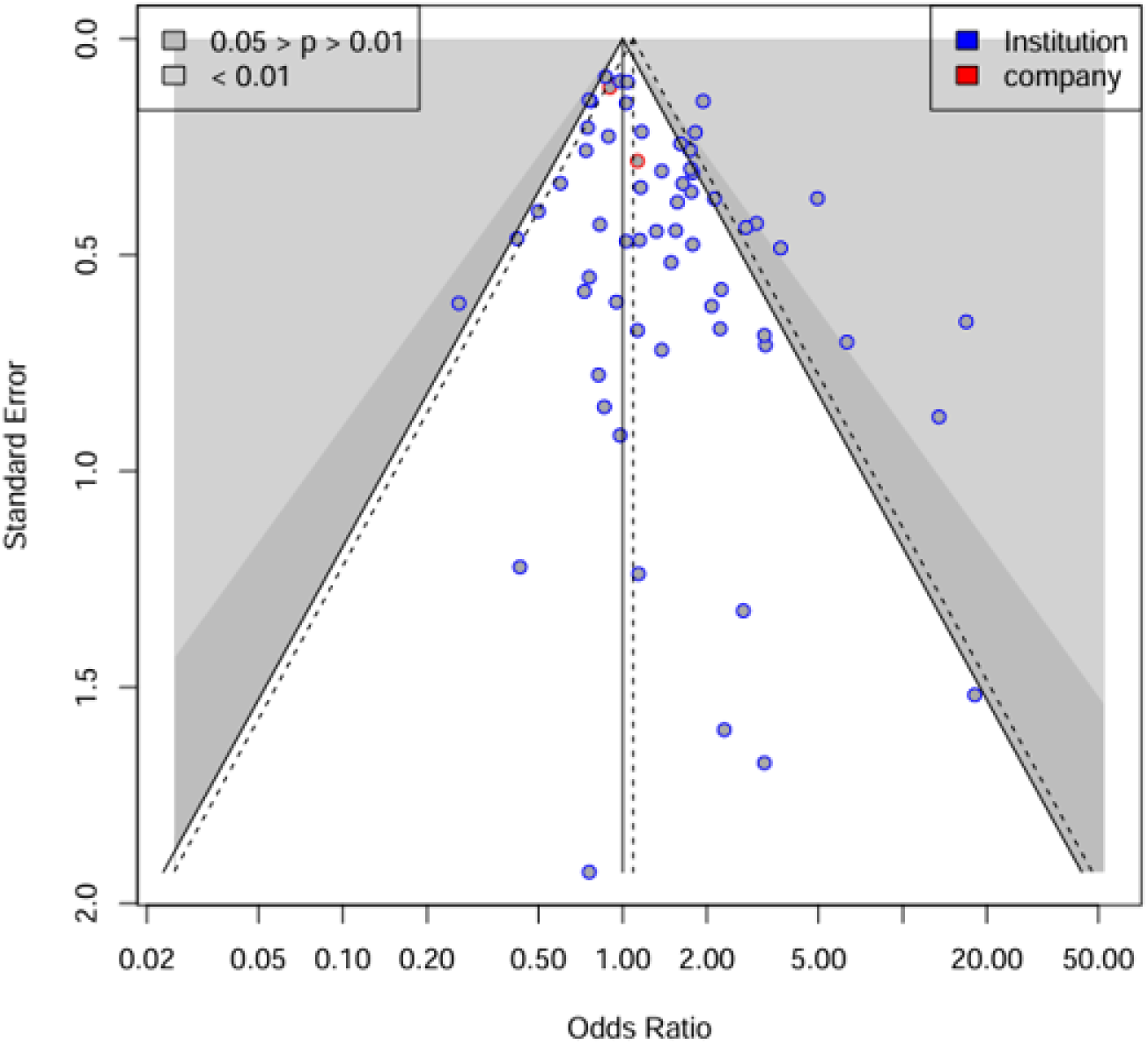
Funnel plot of the association [SLCO1B1 SNPs – Statins – SAMS k = 62] based on sponsor type -. k: number of point estimates. In the funnel plots each point is an estimation of the associations, color corresponds to sponsor type. The white, dark and light grey zones stand for a p value of the odds ratio i) non-significant, ii) between 0.05 and 0.01, and iii) <0.01, respectively. The dashed triangle stands for the estimation of the meta-analysis of the association, without adjusting for a potential publication bias

### Supplementary material 13 (S13): Funnel plot of the association [SLCO1B1 SNPs – Statins – SAMS] based on ethnic group as reported in systematic reviews

**Figure S13.**
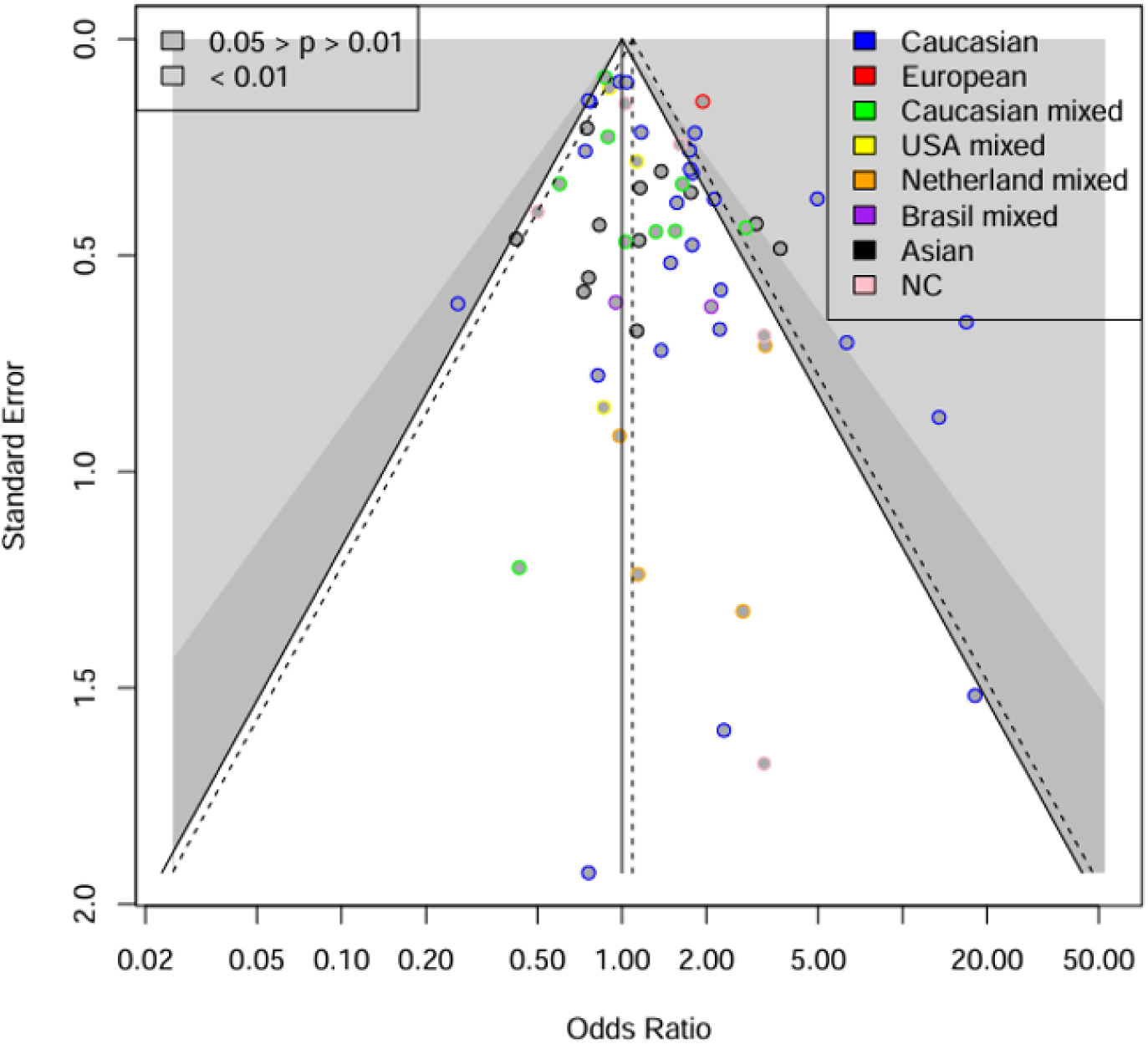
Funnel plot of the association [SLCO1B1 SNPs – Statins – SAMS k = 62] based on ethnic group as reported in clinical studies-. k: number of point estimates. In the funnel plots each point is an estimation of the associations, color corresponds to sponsor type. The white, dark and light grey zones stand for a p value of the odds ratio i) non-significant, ii) between 0.05 and 0.01, and iii) <0.01, respectively. The dashed triangle stands for the estimation of the meta-analysis of the association, without adjusting for a potential publication bias

## References

[1] Sacks FM, Pfeffer MA, Moye LA, Rouleau JL, Rutherford JD, Cole TG, et al. The Effect of Pravastatin on Coronary Events after Myocardial Infarction in Patients with Average Cholesterol Levels. N Engl J Med 1996;335:1001–9. 10.1056/NEJM199610033351401.

[2] Heart Protection Study Collaborative Group. MRC/BHF Heart Protection Study of cholesterol lowering with simvastatin in 20,536 high-risk individuals: a randomised placebo-controlled trial. Lancet Lond Engl 2002;360:7–22. 10.1016/S0140-6736(02)09327-3.

[3] Ridker PM, Danielson E, Fonseca FAH, Genest J, Gotto AM, Kastelein JJP, et al. Rosuvastatin to Prevent Vascular Events in Men and Women with Elevated C-Reactive Protein. N Engl J Med 2008;359:2195–207. 10.1056/NEJMoa0807646.

[4] Warden BA, Guyton JR, Kovacs AC, Durham JA, Jones LK, Dixon DL, et al. Assessment and management of statin-associated muscle symptoms (SAMS): A clinical perspective from the National Lipid Association. J Clin Lipidol 2023;17:19–39. 10.1016/j.jacl.2022.09.001.

[5] Romaine SPR, Bailey KM, Hall AS, Balmforth AJ. The influence of SLCO1B1 (OATP1B1) gene polymorphisms on response to statin therapy. Pharmacogenomics J 2010;10:1–11. 10.1038/tpj.2009.54.

[6] Link L. SLCO1B1 Variants and Statin-Induced Myopathy — A Genomewide Study. N Engl J Med 2008;359:789–99. 10.1056/NEJMoa0801936.

[7] Brunham LR, Lansberg PJ, Zhang L, Miao F, Carter C, Hovingh GK, et al. Differential effect of the rs4149056 variant in SLCO1B1 on myopathy associated with simvastatin and atorvastatin. Pharmacogenomics J 2012;12:233–7. 10.1038/tpj.2010.92.

[8] Voora D, Shah S, Spasojevic I, Ali S, Reed C, Salisbury B, et al. The SLCO1B1*5 genetic variant is associated with statin-induced side effects. J Am Coll Cardiol 2009;54:1609–1616. 10.1016/j.jacc.2009.04.053.

[9] Carr DF, O’Meara H, Jorgensen AL, Campbell J, Hobbs M, McCann G, et al. SLCO1B1 genetic variant associated with statin-induced myopathy: a proof-of-concept study using the clinical practice research datalink. Clin Pharmacol Ther 2013;94:695–701. 10.1038/clpt.2013.161.

[10] Danik JS, Chasman DI, MacFadyen JG, Nyberg F, Barratt BJ, Ridker PM. Lack of association between SLCO1B1 polymorphisms and clinical myalgia following rosuvastatin therapy. Pharmacoepidemiol Drug Saf 2012;21:185–6. 10.1002/pds.3324.

[11] Puccetti L, Ciani F, Auteri A. Genetic involvement in statins induced myopathy. Preliminary data from an observational case-control study. Atherosclerosis 2010;211:28–9. 10.1016/j.atherosclerosis.2010.02.026.

[12] Cooper-DeHoff RM, Niemi M, Ramsey LB, Luzum JA, Tarkiainen EK, Straka RJ, et al. The Clinical Pharmacogenetics Implementation Consortium Guideline for SLCO1B1, ABCG2, and CYP2C9 genotypes and Statin-Associated Musculoskeletal Symptoms. Clin Pharmacol Ther 2022;111:1007–21. 10.1002/cpt.2557.

[13] Obeng AO, Scott SA, Kaszemacher T, Ellis SB, Mejia A, Gomez A, et al. Prescriber Adoption of SLCO1B1 Genotype-Guided Simvastatin Clinical Decision Support in a Clinical Pharmacogenetics Program. Clin Pharmacol Ther 2023;113:321–7. 10.1002/cpt.2773.

[14] Turner EH, Matthews AM, Linardatos E, Tell RA, Rosenthal R. Selective Publication of Antidepressant Trials and Its Influence on Apparent Efficacy. N Engl J Med 2008;358:252–60. 10.1056/NEJMsa065779.

[15] Wang C-H, Li C-H, Hsieh R, Fan C-Y, Hsu T-C, Chang W-C, et al. Proton pump inhibitors therapy and the risk of pneumonia: a systematic review and meta-analysis of randomized controlled trials and observational studies. Expert Opin Drug Saf 2019;18:163–72. 10.1080/14740338.2019.1577820.

[16] Pan Z, Trikalinos TA, Kavvoura FK, Lau J, Ioannidis JPA. Local literature bias in genetic epidemiology: an empirical evaluation of the Chinese literature. PLoS Med 2005;2:e334. 10.1371/journal.pmed.0020334.

[17] Bally S, Cottin J, Gagnieu MC, Lega JC, Verstuyft C, Rheims S, et al. Publication bias in pharmacogenetics of adverse reaction to antiseizure drugs: An umbrella review and a meta- epidemiological study. PloS One 2022;17:e0278839. 10.1371/journal.pone.0278839.

[18] Jiang J, Tang Q, Feng J, Dai R, Wang Y, Yang Y, et al. Association between SLCO1B1 −521T>C and −388A>G polymorphisms and risk of statin-induced adverse drug reactions: A meta-analysis. SpringerPlus 2016;5:1368. 10.1186/s40064-016-2912-z.

[19] Xiang Q, Chen S-Q, Ma L-Y, Hu K, Zhang Z, Mu G-Y, et al. Association between SLCO1B1 T521C polymorphism and risk of statin-induced myopathy: a meta-analysis. Pharmacogenomics J 2018;18:721–9. 10.1038/s41397-018-0054-0.

[20] Duval S, Tweedie R. Trim and Fill: A Simple Funnel-Plot–Based Method of Testing and Adjusting for Publication Bias in Meta-Analysis. Biometrics 2000;56:455–63. 10.1111/j.0006-341X.2000.00455.x.

[21] Higgins JPT, Thomas J, Chandler J, Cumpston M, Li T, Page MJ, Welch VA (editors). Cochrane Handbook for Systematic Reviews of Interventions version 6.3 (updated February 2022). Cochrane, 2022. Available from www.training.cochrane.org/handbook. n.d.

[22] Moher D, Shamseer L, Clarke M, Ghersi D, Liberati A, Petticrew M, et al. Preferred reporting items for systematic review and meta-analysis protocols (PRISMA-P) 2015 statement. Syst Rev 2015;4:1. 10.1186/2046-4053-4-1.

[23] Krnic Martinic M, Pieper D, Glatt A, Puljak L. Definition of a systematic review used in overviews of systematic reviews, meta-epidemiological studies and textbooks. BMC Med Res Methodol 2019;19:1–12. 10.1186/s12874-019-0855-0.

[24] Egger M, Smith GD, Schneider M, Minder C. Bias in meta-analysis detected by a simple, graphical test. BMJ 1997;315:629–34. 10.1136/bmj.315.7109.629.

[25] Gombault C, Grenet G, Segurel L, Duret L, Gueyffier F, Cathébras P, et al. Population designations in biomedical research: Limitations and perspectives. HLA 2023;101:3–15. 10.1111/tan.14852.

[26] Warrens MJ. Inequalities between multi-rater kappas. Adv Data Anal Classif 2010;4:271–86. 10.1007/s11634-010-0073-4.

[27] Begg CB, Mazumdar M. Operating characteristics of a rank correlation test for publication bias. Biometrics 1994;50:1088–101.

[28] Bartoš F, Maier M, Wagenmakers E, Doucouliagos H, Stanley TD. Robust Bayesian meta- analysis: Model-averaging across complementary publication bias adjustment methods. Res Synth Methods 2023;14:99–116. 10.1002/jrsm.1594.

[29] Higgins JPT, Thompson SG, Deeks JJ, Altman DG. Measuring inconsistency in meta-analyses. BMJ 2003;327:557–60. 10.1136/bmj.327.7414.557.

[30] Maier M, VanderWeele TJ, Mathur MB. Using selection models to assess sensitivity to publication bias: A tutorial and call for more routine use. Campbell Syst Rev 2022;18:e1256. 10.1002/cl2.1256.

[31] Nobre A de P, de Melo GM, Shanks DR. Publication bias casts doubt on implicit processing in inattentional blindness. Neurosci Biobehav Rev 2022;140:104775. 10.1016/j.neubiorev.2022.104775.

[32] Holgado D, Mesquida C, Román-Caballero R. Assessing the Evidential Value of Mental Fatigue and Exercise Research. Sports Med Auckl NZ 2023. 10.1007/s40279-023-01926-w.

[33] Borawski J, Papadatou-Pastou M, Packheiser J, Ocklenburg S. Handedness in post-traumatic stress disorder: A meta-analysis. Neurosci Biobehav Rev 2023;145:105009. 10.1016/j.neubiorev.2022.105009.

[34] R Core Team (2022). R: A language and environment for statistical computing. R Foundation for Statistical Computing, Vienna, Austria. URL https://www.R-project.org/. n.d.

[35] Balduzzi S, Rücker G, Schwarzer G. How to perform a meta-analysis with R: a practical tutorial. Evid Based Ment Health 2019;22:153–60. 10.1136/ebmental-2019-300117.

[36] Hou Q, Li S, Li L, Li Y, Sun X, Tian H. Association Between SLCO1B1 Gene T521C Polymorphism and Statin-Related Myopathy Risk: A Meta-Analysis of Case-Control Studies. Medicine (Baltimore) 2015;94:e1268. 10.1097/MD.0000000000001268.

[37] de Keyser CE, Peters BJM, Becker ML, Visser LE, Uitterlinden AG, Klungel OH, et al. The SLCO1B1 c.521T>C polymorphism is associated with dose decrease or switching during statin therapy in the Rotterdam Study. Pharmacogenet Genomics 2014;24:43. 10.1097/FPC.0000000000000018.

[38] Nguyen KA, Li L, Lu D, Yazdanparast A, Wang L, Kreutz RP, et al. A comprehensive review and meta-analysis of risk factors for statin-induced myopathy. Eur J Clin Pharmacol 2018;74:1099–109. 10.1007/s00228-018-2482-9.

[39] Turongkaravee S, Jittikoon J, Lukkunaprasit T, Sangroongruangsri S, Chaikledkaew U, Thakkinstian A. A systematic review and meta-analysis of genotype-based and individualized data analysis of SLCO1B1 gene and statin-induced myopathy. Pharmacogenomics J 2021;21:296–307. 10.1038/s41397-021-00208-w.

[40] Du Y, Wang S, Chen Z, Sun S, Zhao Z, Li X. Association of SLCO1B1 Polymorphisms and Atorvastatin Safety and Efficacy: A Meta-analysis. Curr Pharm Des 2018;24:4044–50. 10.2174/1381612825666181219163534.

[41] Xiang Q, Zhang X-D, Mu G-Y, Wang Z, Liu Z-Y, Xie Q-F, et al. Correlation between single- nucleotide polymorphisms and statin-induced myopathy: a mixed-effects model meta-analysis. Eur J Clin Pharmacol 2021;77:569–81. 10.1007/s00228-020-03029-1.

[42] Ioannidis JPA. Why Most Published Research Findings Are False. PLoS Med 2005;2:e124. 10.1371/journal.pmed.0020124.

[43] Ioannidis JPA, Trikalinos TA. The appropriateness of asymmetry tests for publication bias in meta-analyses: a large survey. CMAJ Can Med Assoc J 2007;176:1091–6. 10.1503/cmaj.060410.

[44] Wood L, Egger M, Gluud LL, Schulz KF, Jüni P, Altman DG, et al. Empirical evidence of bias in treatment effect estimates in controlled trials with different interventions and outcomes: meta- epidemiological study. BMJ 2008;336:601. 10.1136/bmj.39465.451748.AD.

